# The health beliefs, attitudes, and intentions of males toward pregnancy planning and preconception health and care: a systematic review

**DOI:** 10.1101/2025.03.25.25324432

**Authors:** Tristan Carter, Danielle Schoenaker, Jon Adams, Amie Steel

**Affiliations:** School of Public Health, Faculty of Health, University of Technology Sydney, Sydney, Australia, 2006; School of Human Development and Health, Faculty of Medicine, University of Southampton, Southampton, UK; MRC Lifecourse Epidemiology Centre, University of Southampton, Southampton, UK; NIHR Southampton Biomedical Research Centre, University of Southampton and University Hospital Southampton NHS Foundation Trust, Southampton, United Kingdom

**Keywords:** Preconception, paternal, health beliefs, attitudes, intentions

## Abstract

**Introduction:** The preconception period is an opportunity to address health-related behaviours to optimise pregnancy and child health outcomes. However, preconception health research and practice are primarily focused on females while similar attention on males remains underdeveloped. To address evidence gaps and inform effective paternal preconception health support, the aim of this systematic review was to identify the health beliefs, attitudes, and intentions of males toward pregnancy planning and preconception health and care.

**Methods:** A literature search was conducted in seven databases: Medline, Embase, PubMed, CINAHL, PsycINFO, Scopus, and Web of Science to identify original research regarding pregnancy planning or preconception health beliefs, attitudes, and/or intentions among generally healthy adult males. Methodological rigour of included studies was assessed using the Newcastle Ottawa Scale (NOS) and the Critical Appraisal Skills Programme (CASP) Qualitative Studies checklist.

**Results:** Nine studies were included; cross sectional studies (n=6); a qualitative exploratory case study (n=1); a mixed method study (n=1) and a study incorporating qualitative and quantitative surveys (n=1). Analysis identified three broad themes: 1) *Importance of Preconception Health and Care*; 2) *Paternal Preconception Behaviours*; and 3) *Inequalities in Preconception Health and Preconception Ca*.*re* Findings reveal 1) Many males did not attend a preconception care consultation and believed it was not needed, or they already knew enough about a healthy pregnancy. 2) Males often agreed that smoking and alcohol consumption can affect the quality of their sperm and sometimes agreed it is important to consume a healthy preconception diet and to be physically active to achieve a healthy weight before conception. 3) For many males, there was a tendency to direct a greater level of responsibility to the female than to themselves regarding preconception health. African American males can feel marginalised.

**Conclusion:** Males do not always opt for a preconception consultation and many believe they are adequately prepared for a healthy pregnancy. Further, many males place a greater level of responsibility for planning and preparing for pregnancy on their partners rather than themselves. Further research focused upon male experiences and perspectives around preconception health is needed to inform targeted preconception health education, policy and care.

## Introduction

An individual’s health before conception - preconception health - can be optimised through the provision of biomedical, behavioural, and social health care interventions which aim to improve pregnancy and offspring outcomes in the short and long term (1). The preconception period is considered within three separate perspectives; 1) throughout the days to weeks before conception (biological); 2) from the time of conscious intention to conceive – typically weeks to months before conception (individual), and 3) through an individual’s life-course (public health) (2). For an individual or couple with the intention to conceive, the process of pregnancy planning is an opportunity to address health related issues such as bodyweight and health behaviours including diet, smoking, alcohol consumption, recreational drug use, and physical activity to enhance overall health for future generations (3). However, a gender imbalance exists with preconception health research and practice primarily targeting males underdeveloped (4, 5). The available evidence detailing the health behaviours of males during preconception, such as smoking or excessive alcohol consumption, suggest paternal behaviours can have a direct influence on pregnancy and offspring health (6). To inform how males can be supported to make healthy behaviour changes during the preconception period and while planning pregnancy, it is important to firstly understand their beliefs, attitudes, and intentions towards preconception health and care and pregnancy planning.

The belief-behaviour relationship, as defined in the theory of planned behaviour (7), proposes that beliefs towards a behaviour (behavioural beliefs) collate to interact attitudes toward the behaviour and in turn contribute toward the willingness to conduct the consequent behaviour (intention) (7). The belief-behaviour relationship is considered relevant when investigating health behaviours such as physical activity (8) or dietary patterns (9) and may be particularly pertinent when exploring the motivations behind paternal health behaviours to better enable effective care strategies, including the formulation of a reproductive life plan (10). Exploration of paternal beliefs, attitudes, and intentions to engage in preconception care and pregnancy planning is required to address important evidence gaps and overcome potential barriers in the effective support of paternal preconception health behaviours and subsequent behaviour change. Further research must also be conducted investigating paternal beliefs, attitudes, and intentions to engage in preconception care to advance knowledge of the mechanisms which underlie male preconception health behaviours. Understanding the beliefs, attitudes and intentions of males will also support the development of preconception health policy, support public health promotions, and enable health services to formulate effective paternal preconception care services and interventions to initiate and sustain behaviour change. In response, the aim of this review was to identify the health beliefs, attitudes, and intentions of males toward pregnancy planning and preconception health and care.

## Methods

This review was prospectively registered with the International prospective register of systematic reviews - PROSPERO (Registration Number: CRD42023495478) and conducted in accordance with the PRISMA guidelines for reporting systematic reviews (11) and the AMSTAR 2 critical appraisal tool (12).

### Search Strategy

A search of the literature was completed on 3^rd^ January 2024 (see Supplementary File 1 Search Strategy) utilizing the following databases: 1) Medline, 2) Embase, 3) PubMed, 4) CINAHL, 5) PsycINFO, 6) Scopus, and 7) Web of Science. This search was updated on 11^th^ February 2025, with a date limit set since 2024 to capture all recent studies published since the previous search. An initial exploratory search using PubMed and the controlled vocabulary of each database, when available, were used to identify and refine search terms. The final search strategy was developed with an expert health sciences librarian from the University of Technology Sydney (UTS). The Scopus database, and Google Scholar for some papers not in Scopus, were used to further identify all studies which referenced each included study and all studies in the reference list of each included study.

### Selection Criteria

This review included original research manuscripts which report empirical results regarding pregnancy planning or preconception health beliefs, attitudes, and/or intentions specifically for generally healthy adult males. Any study designs (e.g. intervention and observational) and research methods (e.g. quantitative and qualitative) were eligible for inclusion. Papers were included if they identified males’ intention to engage in preconception health behaviour and/or attitudes, beliefs or perspectives to make preconception health behaviour change or engage with preconception care programmes and services. Papers were not included if they only reported on males’ intention to conceive/pregnancy intention (i.e. ‘Do you intend to conceive in the future?’ or ‘Was this a planned pregnancy?’), if they only reported factors which influence intention, or if they only reported fertility or preconception health and care knowledge. These exclusions were applied in the review to remove studies that focused primarily on topics related to a men’s intention to have a child or regarding preconception knowledge which did not provide details of preconception beliefs, attitudes, or intentions or provide context for specific preconception health behaviours such as smoking or alcohol consumption.

Papers excluded were review articles, editorials, commentaries, and letters, which did not report original research, along with articles which did not explore pregnancy planning or preconception health but instead topics such as contraception, abortion, unplanned pregnancy, or ambivalent pregnancy. Also excluded were any papers which included males under 18 years old as the focus of this review is adult males. Males recruited as (health) professionals or medical health experts were also excluded due to their expert understanding of the subject matter and the focus of this review being on the general population. Papers which focused solely on males with a diagnosed illness (e.g. HIV, diabetes) or disability were also excluded as their beliefs and attitudes may be influenced by their conditions.

| Population | Intervention or exposure | Comparator | Outcome |
| --- | --- | --- | --- |
| Adult males aged $\geq 18$ years recruited from the general population. | Beliefs, attitudes, and/or intentions, towards pregnancy planning, preconception health and/or preconception care. | Not required, or as defined in individual studies | Not required |

### Data extraction and synthesis

All studies were imported into Covidence systematic review software (13) and any duplicates were removed automatically. TC screened all papers by title and abstract and then full-text, AS and DS were second independent reviewers for this process and also assisted to resolve conflicts. TC obtained the full-text article for each included paper and, using a data extraction template, performed data extraction on all included papers. Data extraction was also performed by either AS or DS for all papers and extracted data were cross-checked for accuracy and completeness.

The data extracted from each paper included country, study aim, study design, setting, study publication year and the year/s when the study was conducted, participant age and sample size. Details specific for each determinant (i.e. belief, attitude, intention, perspective) were also extracted including the key results and outcomes for each determinant and any relationships between determinants.

The extracted data were initially grouped by exposure as a 1) belief, 2) attitude, or 3) intention. A process of inductive reasoning enabled the research team to determine recurring overarching themes within the extracted data. The paternal preconception beliefs, attitudes, and intentions identified within this review were categorised, tabulated, and reported within three main themes 1) *Importance of Preconception Health and Care*, 2) *Paternal Preconception Behaviours*, and 3) *Inequalities in Preconception Health and Preconception Care*

### Quality assessment

Methodological rigour for studies undertaking quantitative analysis was assessed using the Newcastle Ottawa Scale (NOS) adapted for cross-sectional studies (14). The NOS for cross-sectional studies comprises three domains 1) selection of participants, 2) comparability of study groups, and 3) outcome of interest. A maximum of five stars are allocated for the selection domain [representativeness of the sample, sample size, non-response rate, ascertainment of exposure], a maximum of two stars for the comparability domain [potential confounders], and a maximum of three stars for the outcome domain [assessment of the outcomes and the statistical tests performed]. A total score was allocated out of ten for each study and was then categorized as good (7-10 stars), fair (4-6 stars) or poor quality (0-3 stars) (15).

The Critical Appraisal Skills Programme (CASP) Qualitative Studies checklist was utilized to assess the quality of qualitative studies (16). The CASP checklist is comprised of three sections: Section A-Result validity (Questions 1-6), Section B – What are the results (Questions 7-9), and Section C – Will the results help (Question 10). Each question is scored on a three-point scale (yes, can’t tell, no) except for question 10 which is an open-ended question (how valuable is the research?). Each qualitative study assessed using the CASP reported an affirmative, undecided or negative response for each question which contributed toward the researcher’s final subjective research value by consolidating all the previous questions. For this review, a paper was deemed high value when at least 8 of 10 appraisal criteria were fulfilled and this must include consideration of ethical issues, and a clear statement of the research aims. A paper deemed moderate value affirmed at least six appraisal criteria and included consideration of ethical issues, and a clear statement of the research aims. A paper which was low value did not meet at least six critical appraisal criteria or had at least three “can’t tell” appraisal criteria or did not provide a clear statement of research aims or consider ethical issues.

The quality assessment of each included study was assessed by two independent reviewers and where any conflicts arose, consensus was reached among the reviewers with the assistance of a third reviewer to mediate.

## Results

The search identified 7,762 papers for which titles and abstracts were screened, followed by screening of 172 full-text articles; nine studies were finally included in this review (Figure 1 – PRISMA flow chart). The majority of the papers (n=6) utilised cross sectional surveys (17–22). One study utilised a survey that concurrently captured both quantitative and qualitative survey responses (23), one study was a qualitative exploratory case study design which utilised semi-structured interviews (24), and one study incorporated a mixed methods design comprising a survey and semi-structured interviews (25) (See Table 1). The included studies were conducted in the United States (n=4), United Kingdom (n=1), Canada (n=1), Belgium (n=1), The Netherlands (n=1), and Malaysia (n=1). Males from each study were recruited as a member of a survey panel (n=2), or from the student health clinic of a large public university (n=1) or from a public health clinic (n=1). Recruitment of males also occurred through various other settings including community health centres and public centres for social welfare (n=1), through female partners who participated in the APROPOS-II study (n=1), online via social media (n=2), or from the general public through in person enlistment and recruitment advertisements (n=1). Sample sizes ranged between 15 and 300 participants.

**Figure 1:**
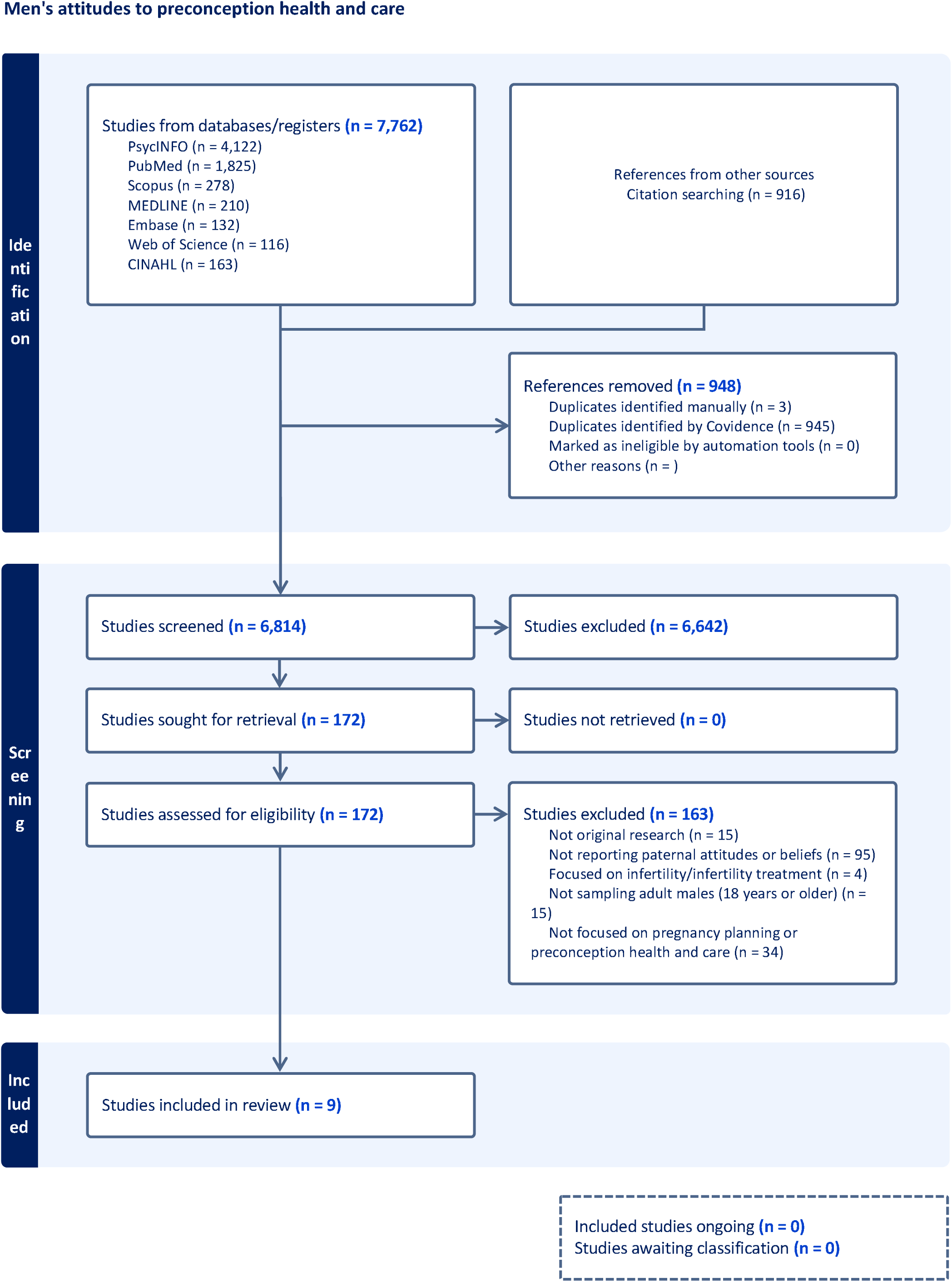
Men’s attitudes to preconception health and care

**Table 1.**
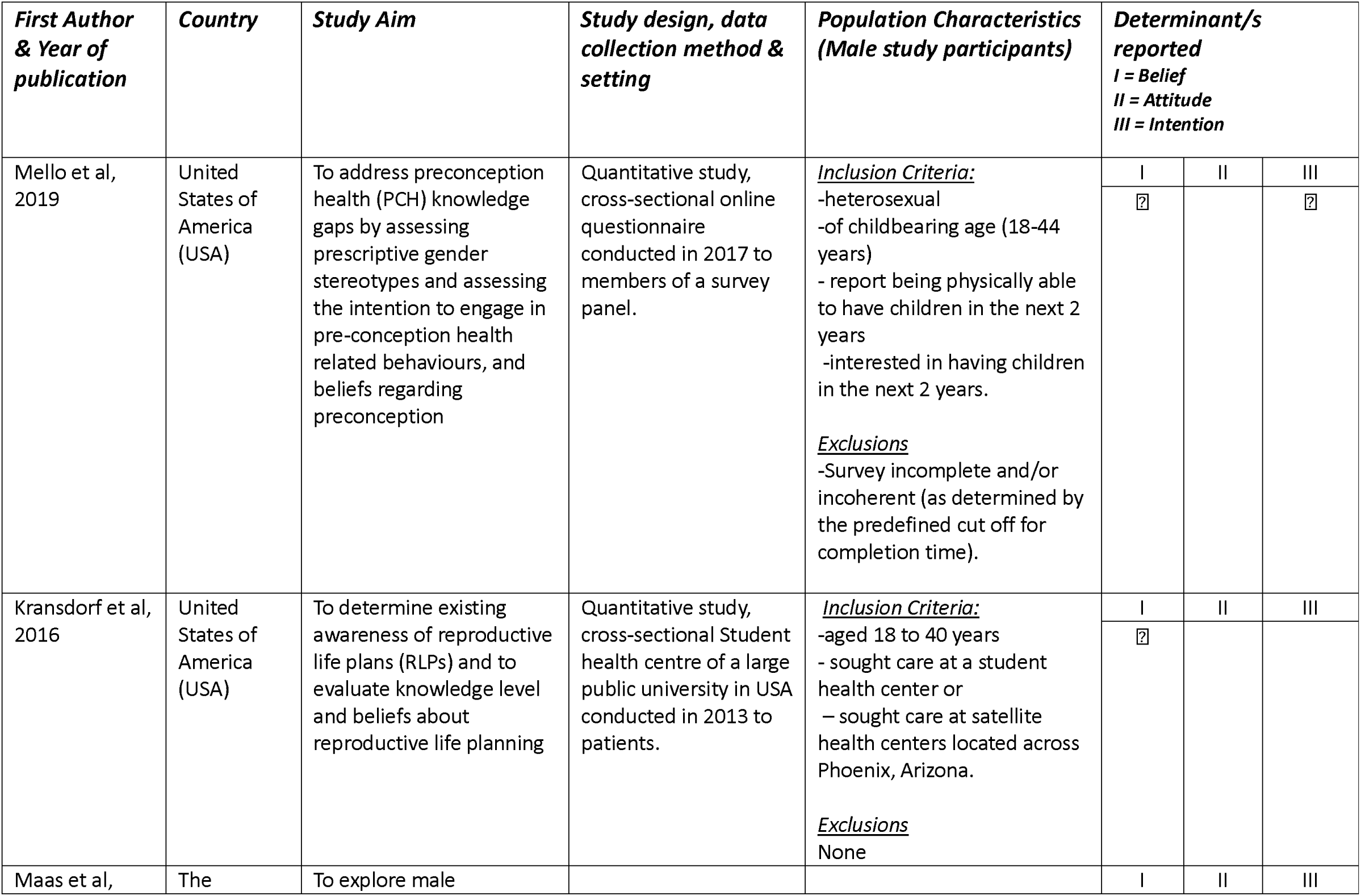
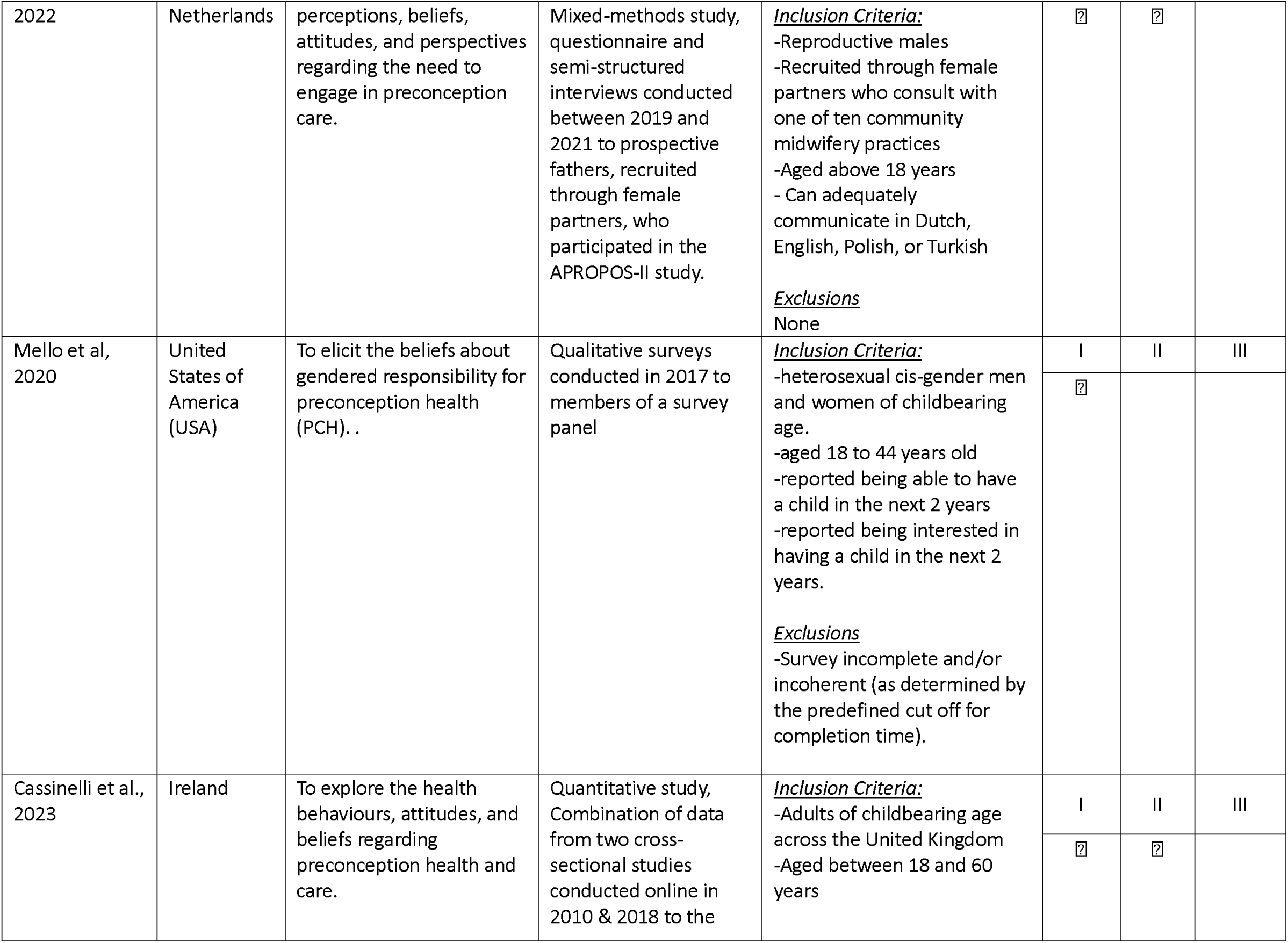
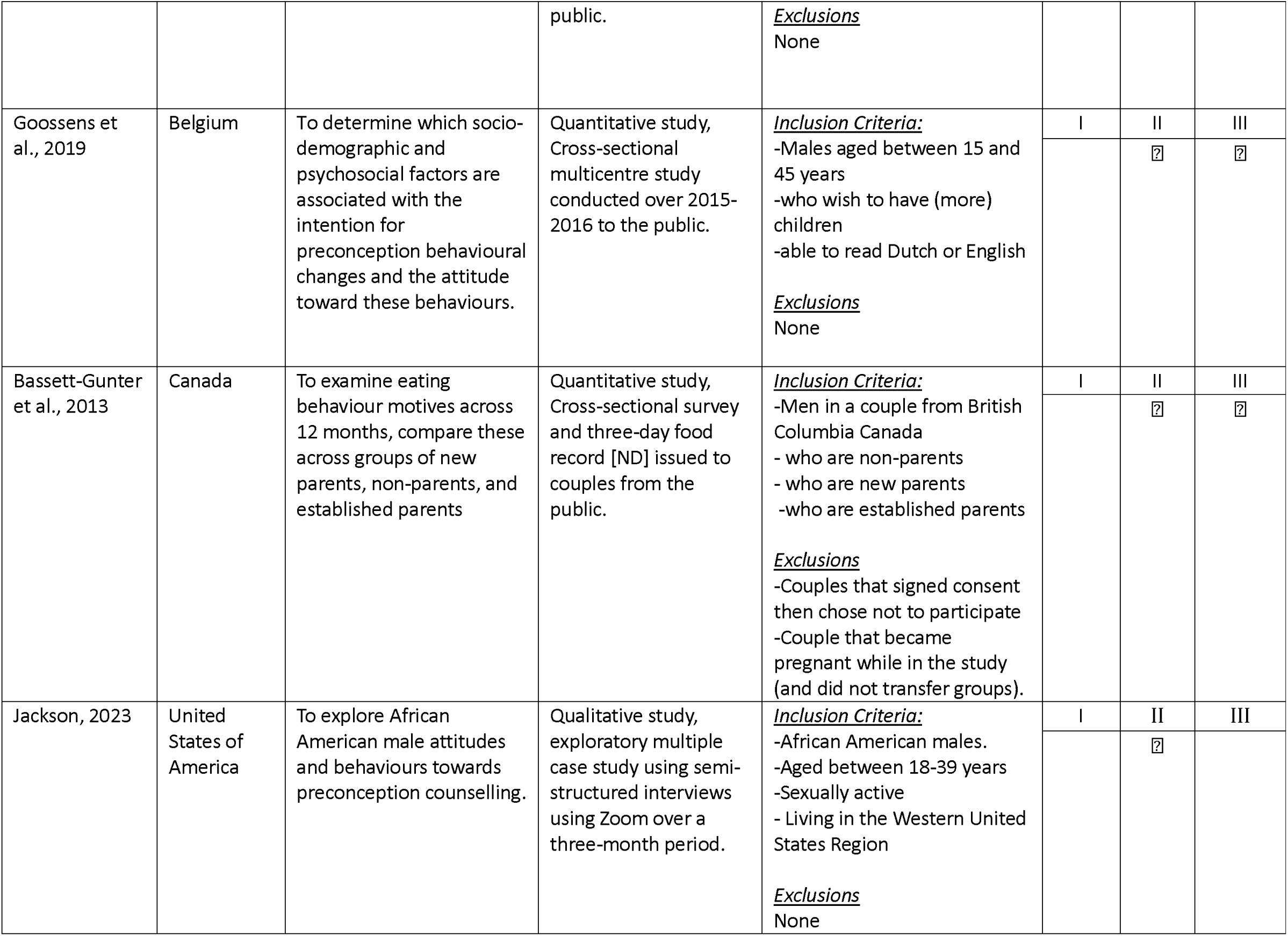
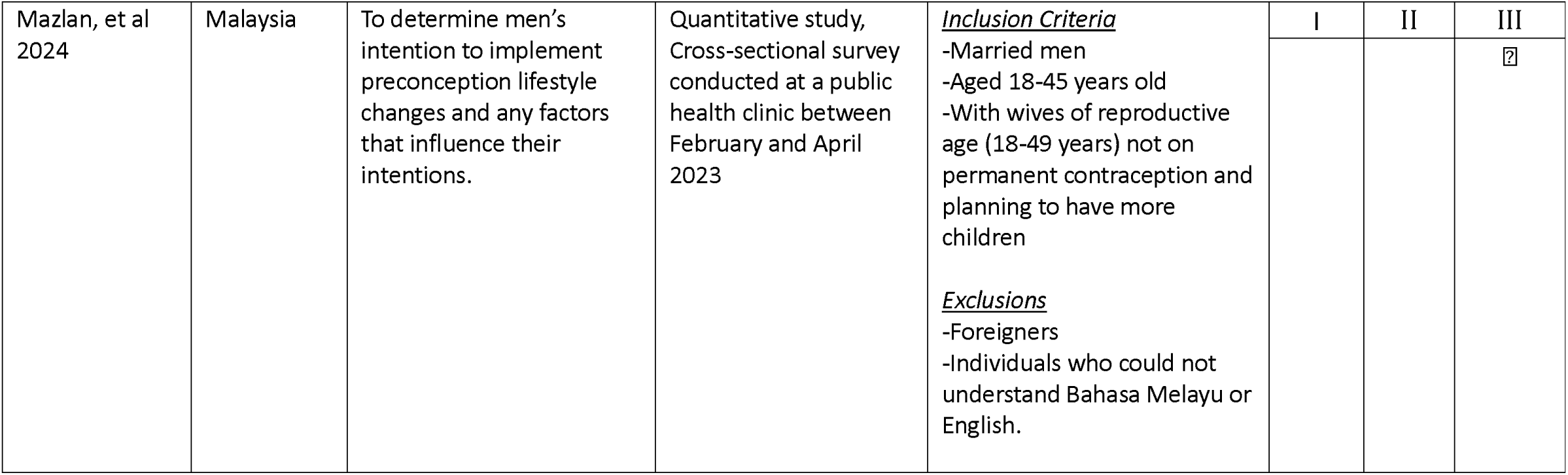
Characteristics of included studies.

### Quality Assessment

Seven studies were assessed for quality using the NOS (17–22, 25) (See Supplementary Table 1) and most (n=6) were determined ‘fair’ quality (17–19, 21, 22, 25), while one was ‘poor’ quality (20) and none were ‘good’ quality. All studies described and used an appropriate statistical test to measure associations and included confidence levels and p-values. None of the studies compared the characteristics between respondents and non-respondents.

The CASP Qualitative Studies checklist was applied to the qualitative conduct and analysis of three studies (23–25) (See Supplementary Table 2). Two studies were classified as high value as they covered at least eight appraisal criteria and had a clear aim, considered ethical issues, justified a research design, and provided adequate data collection and analysis methods (25). The third study was deemed moderate value as it covered only 6 appraisal criteria and did not clearly articulate the research design or the qualitative methodology or provide a clear statement of the findings (23).

Statistically significant and consistently reported results pertaining to the preconception beliefs, attitudes, and intentions identified in this review were categorised into themes and summarised below. All results, including non-significant findings, are presented in Table 2.

**Table 2.**
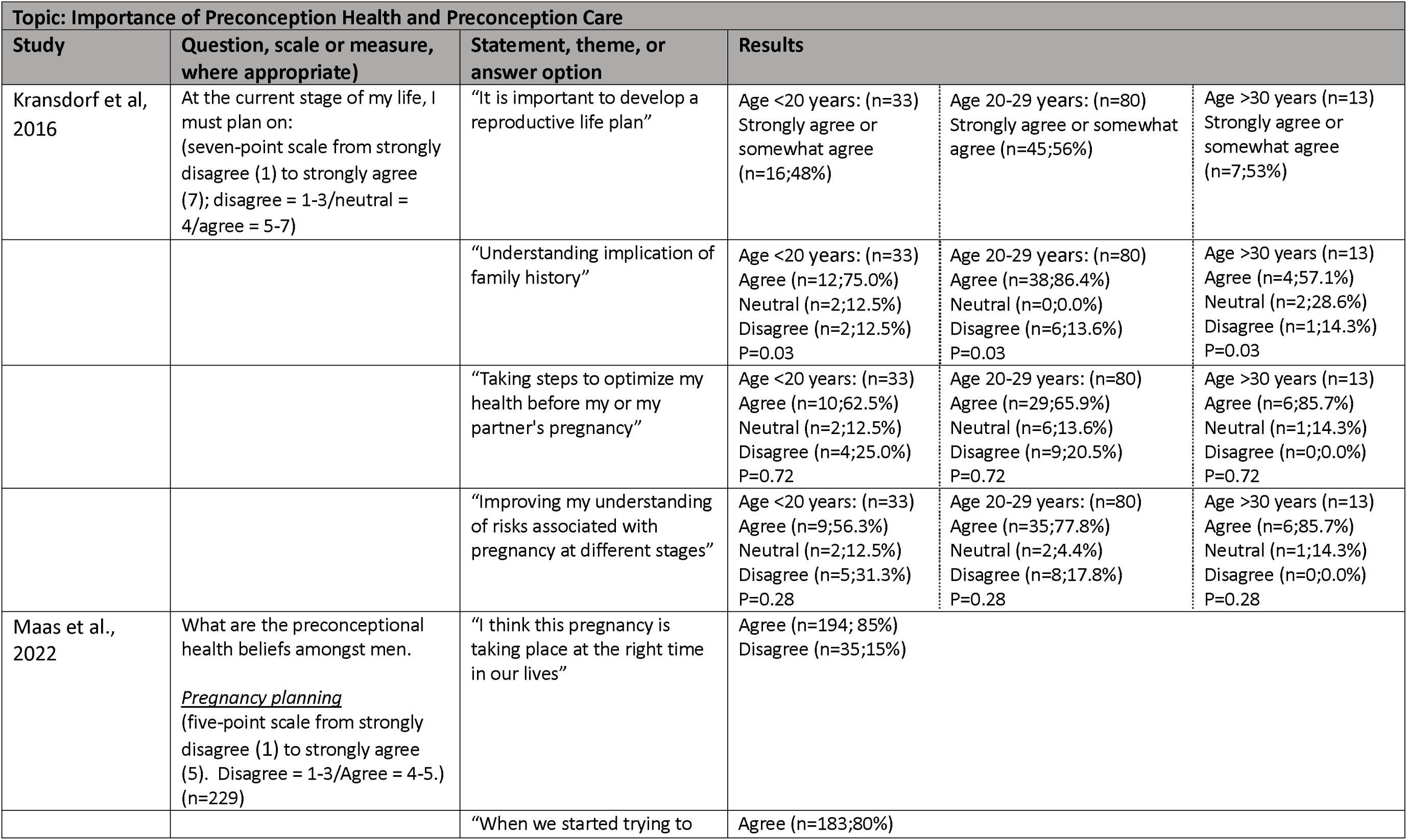
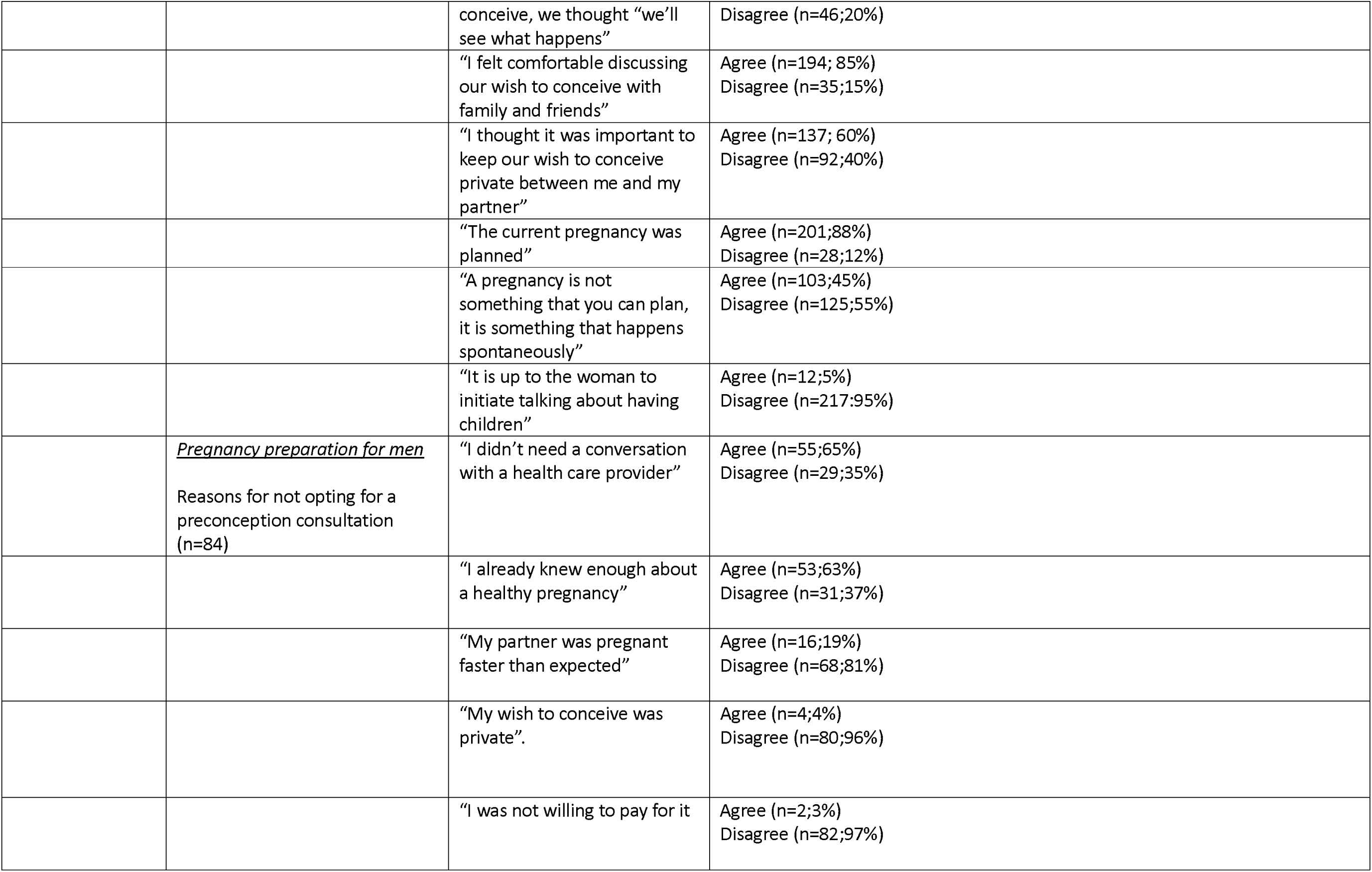
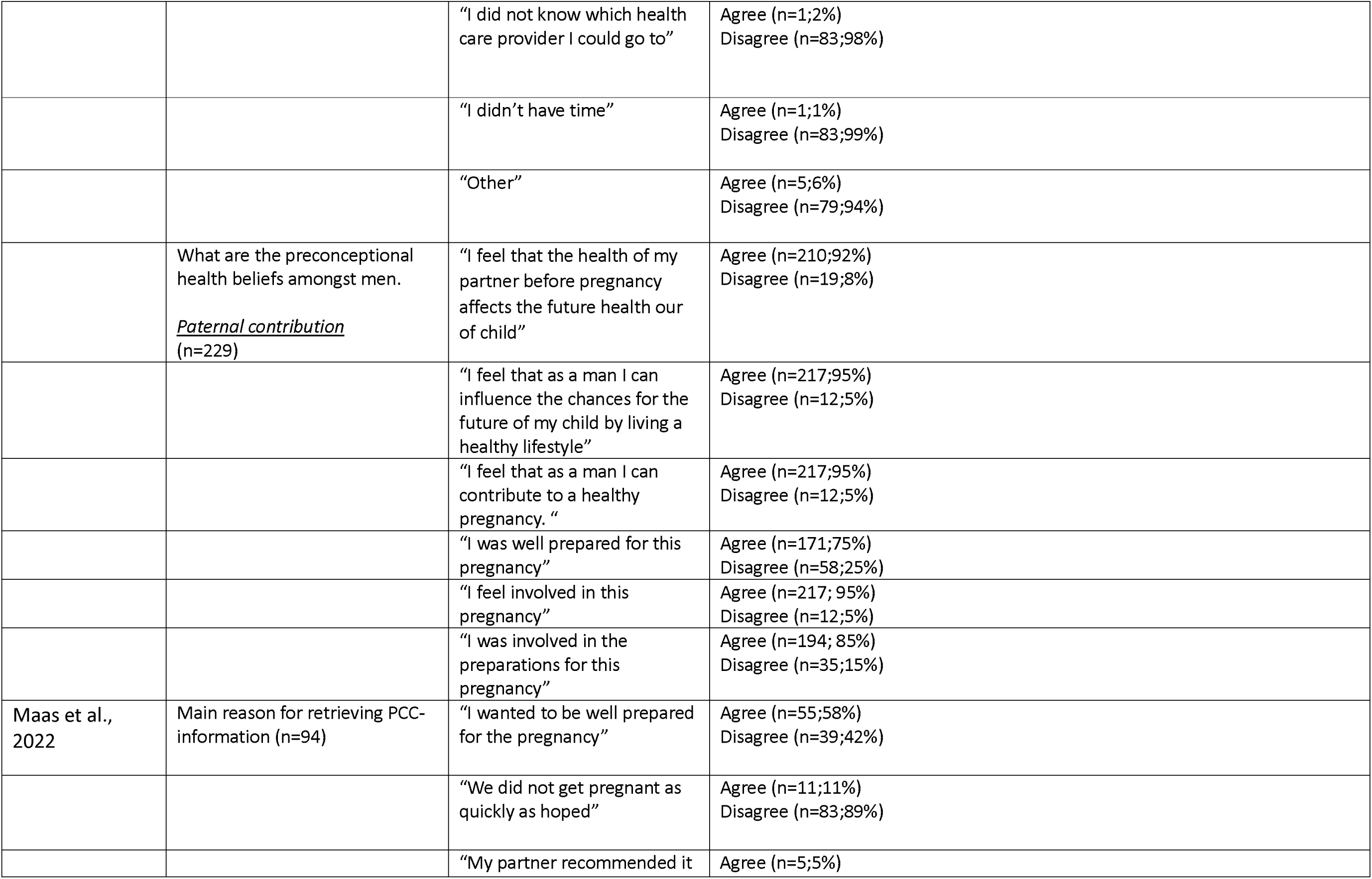
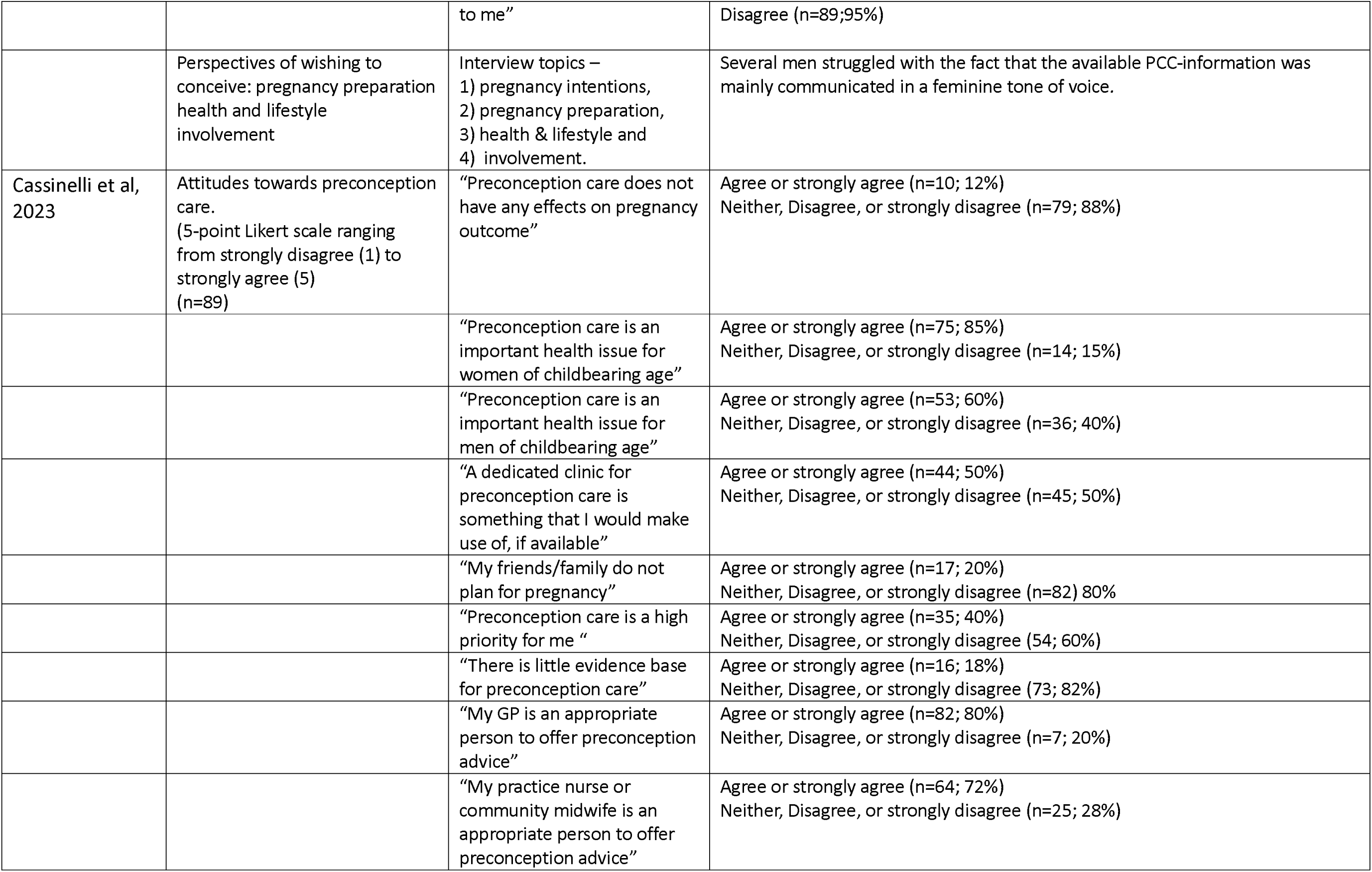
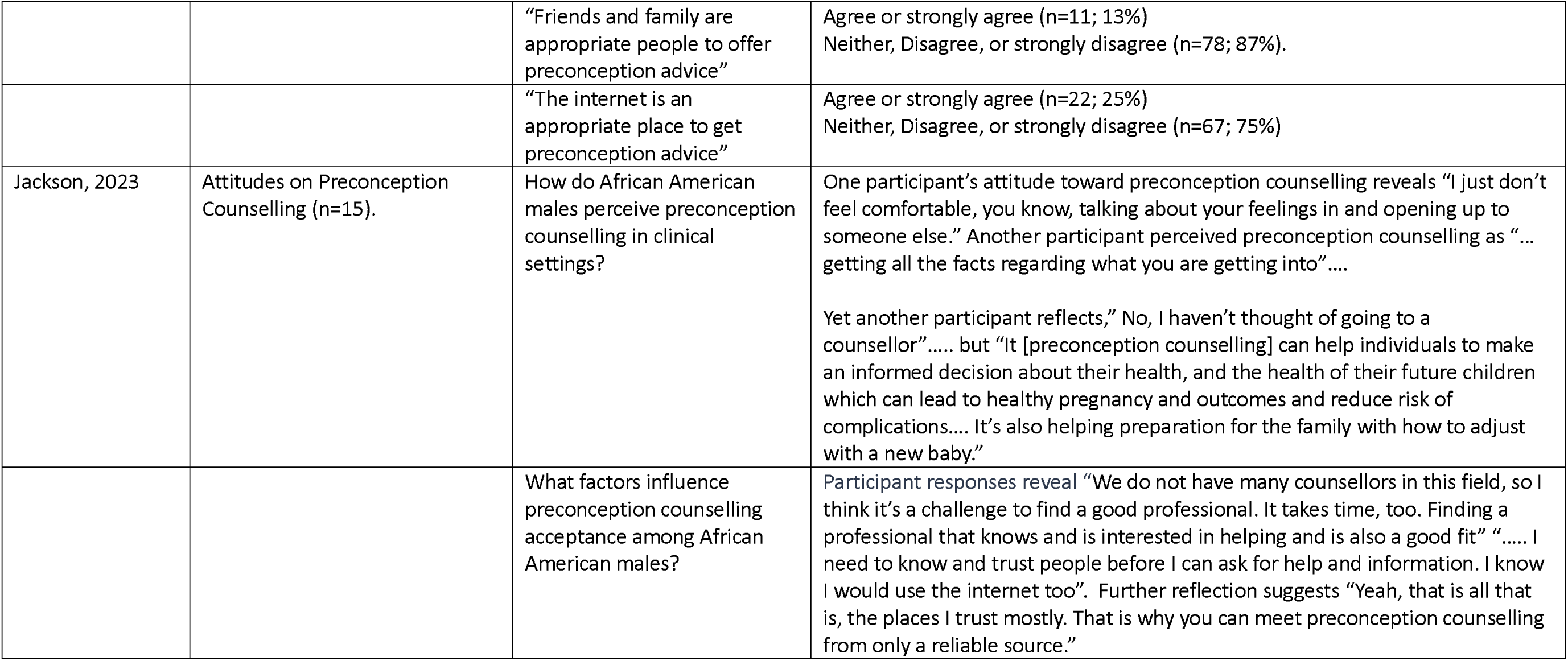
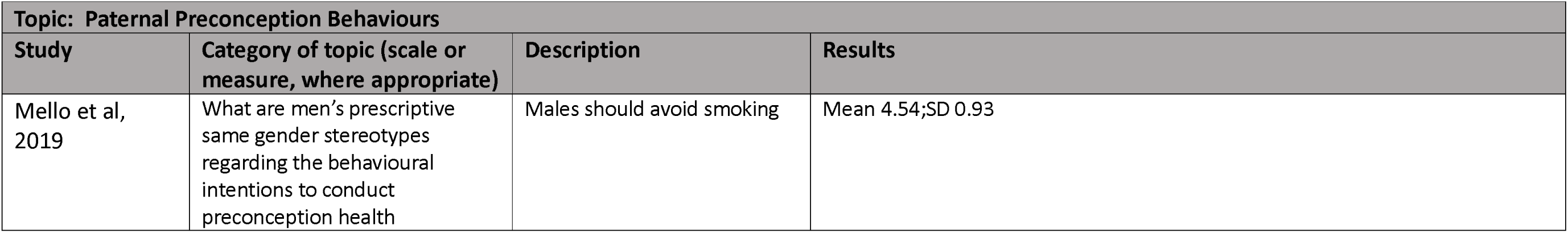
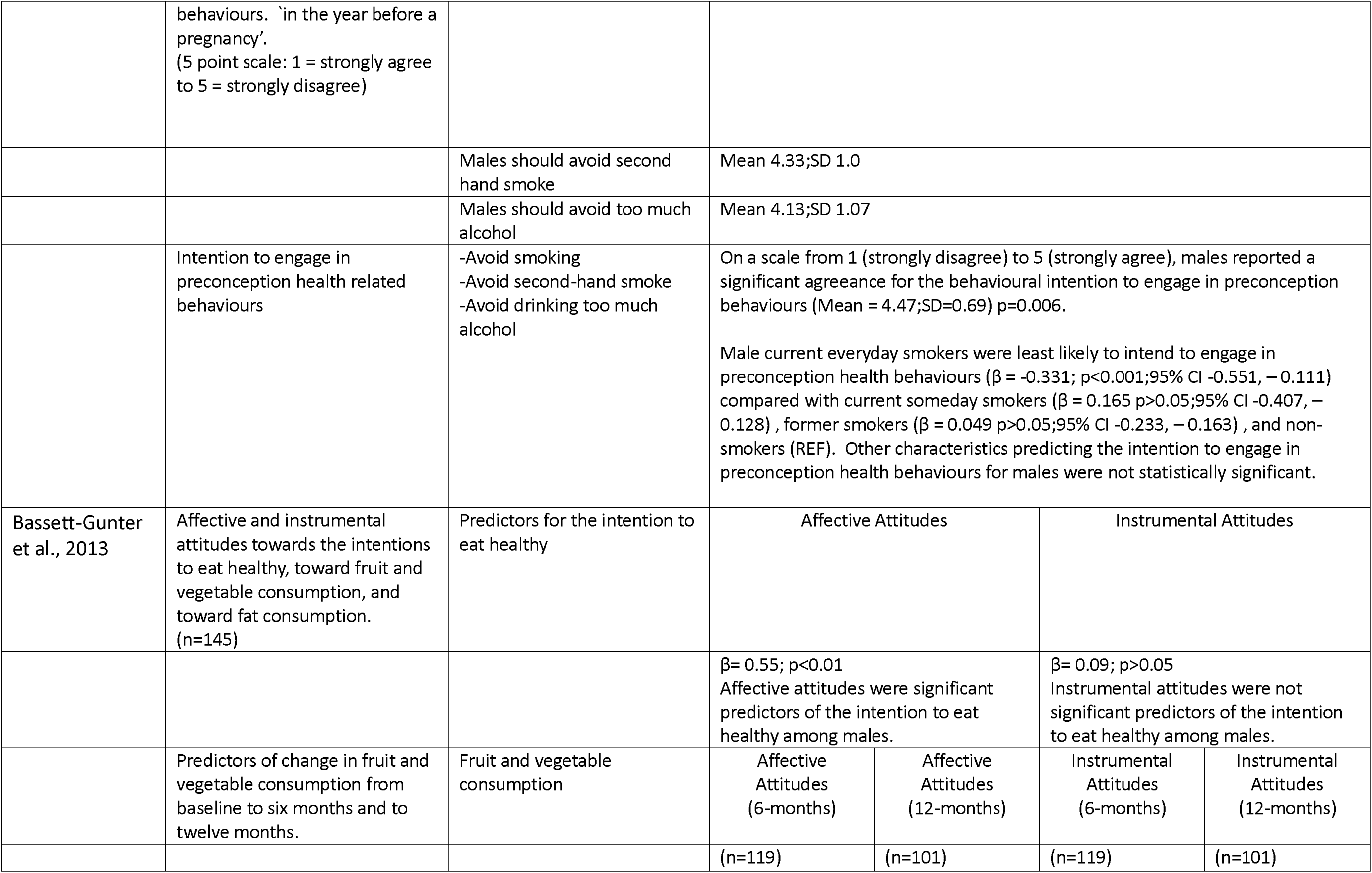
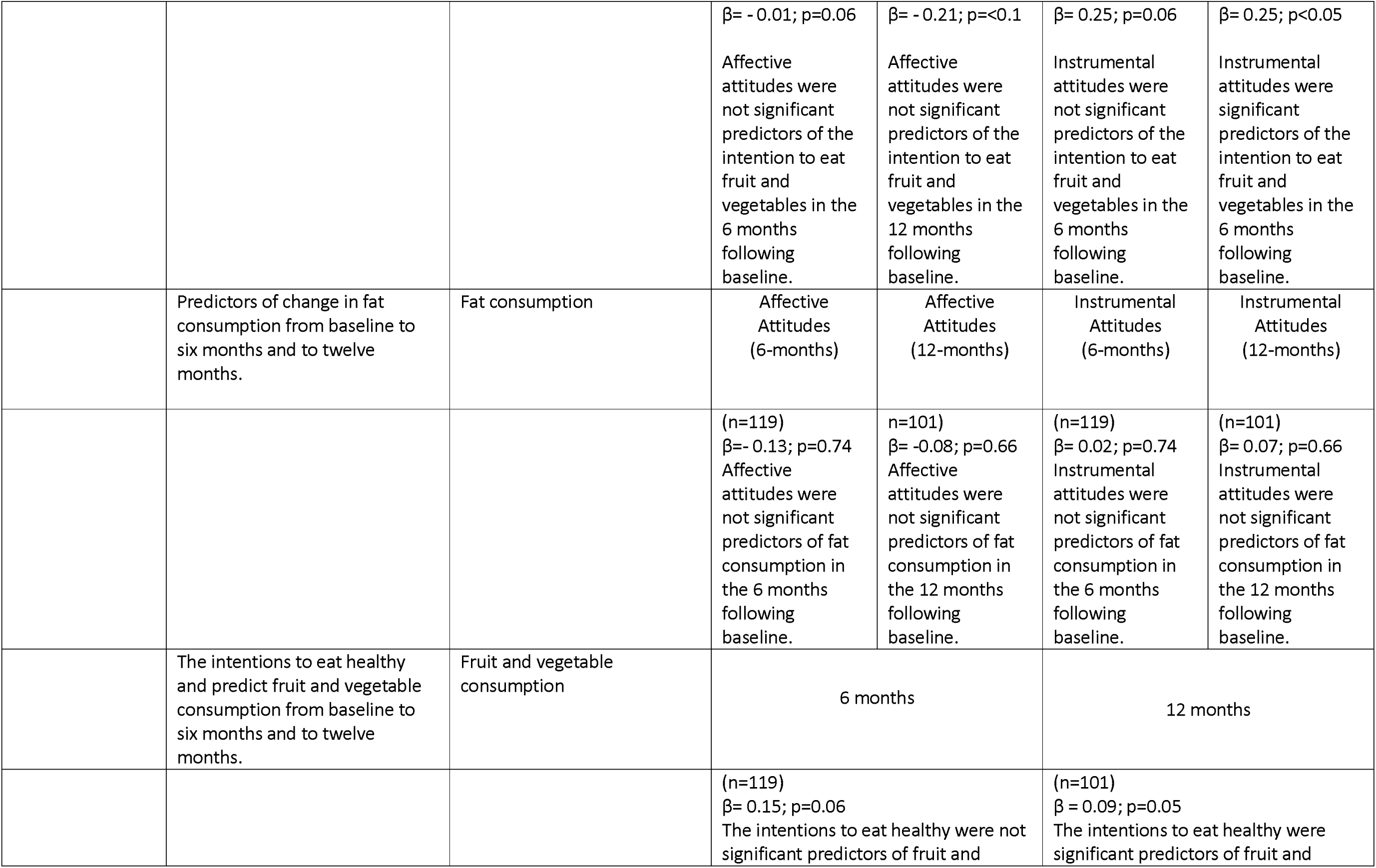
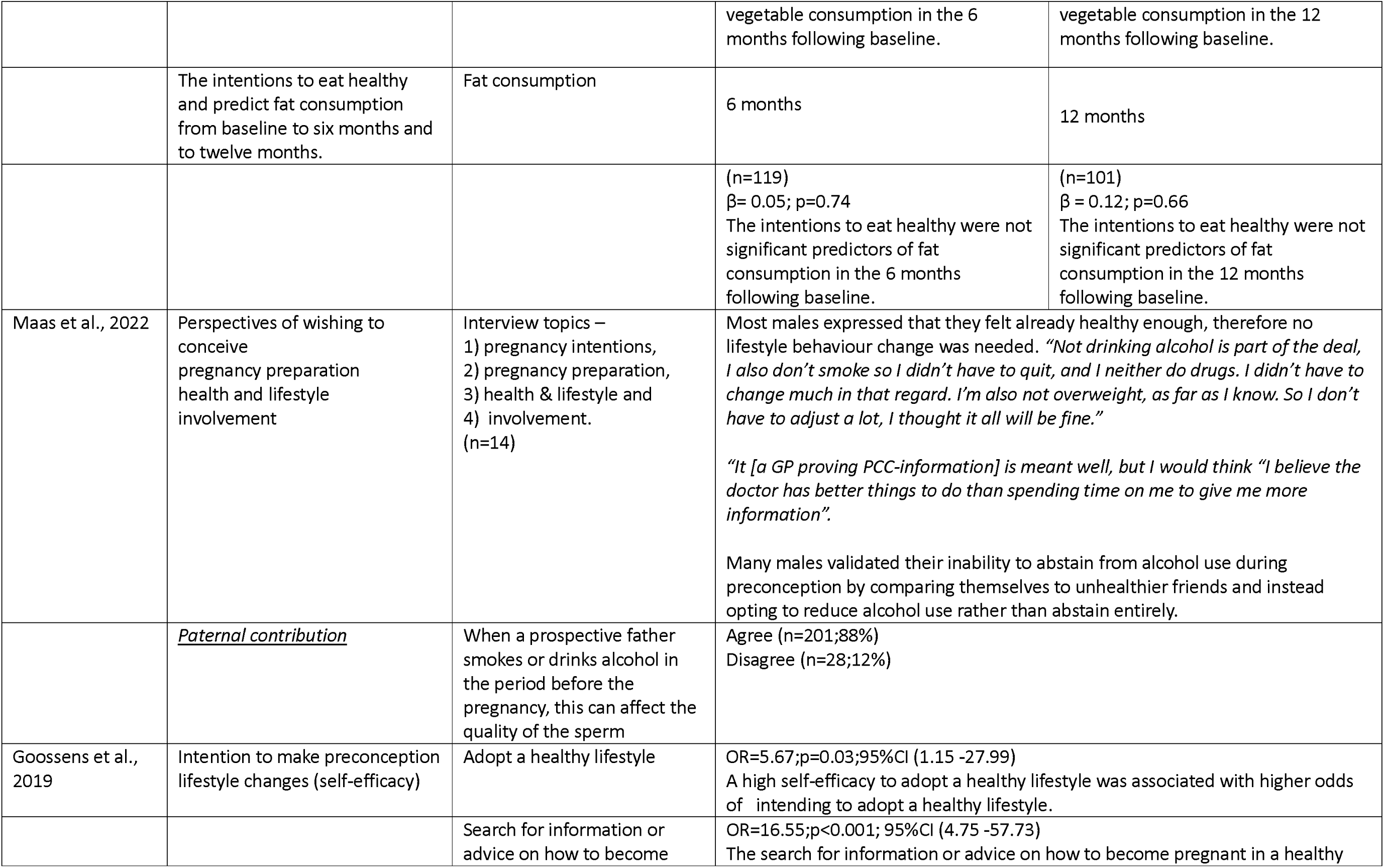
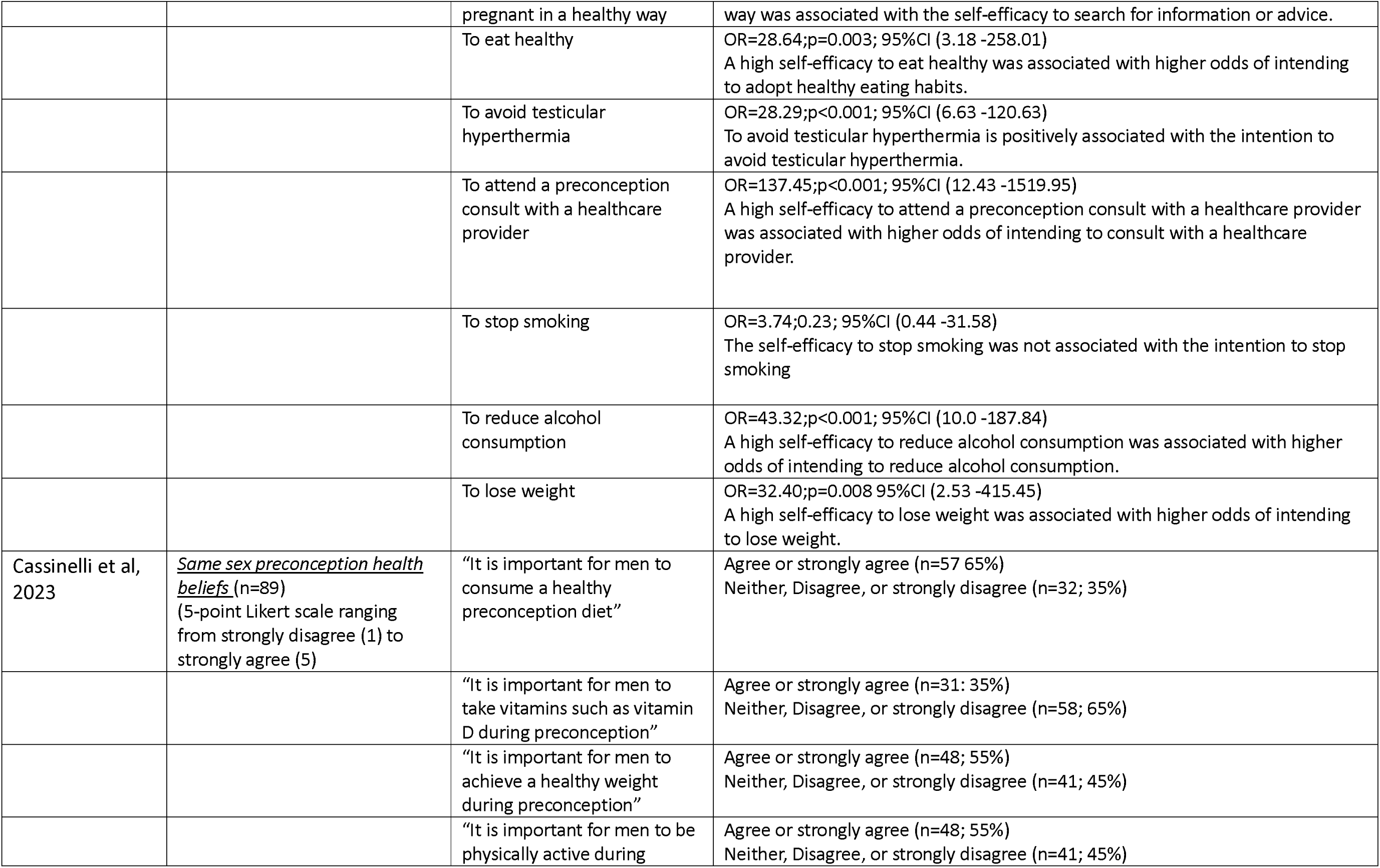
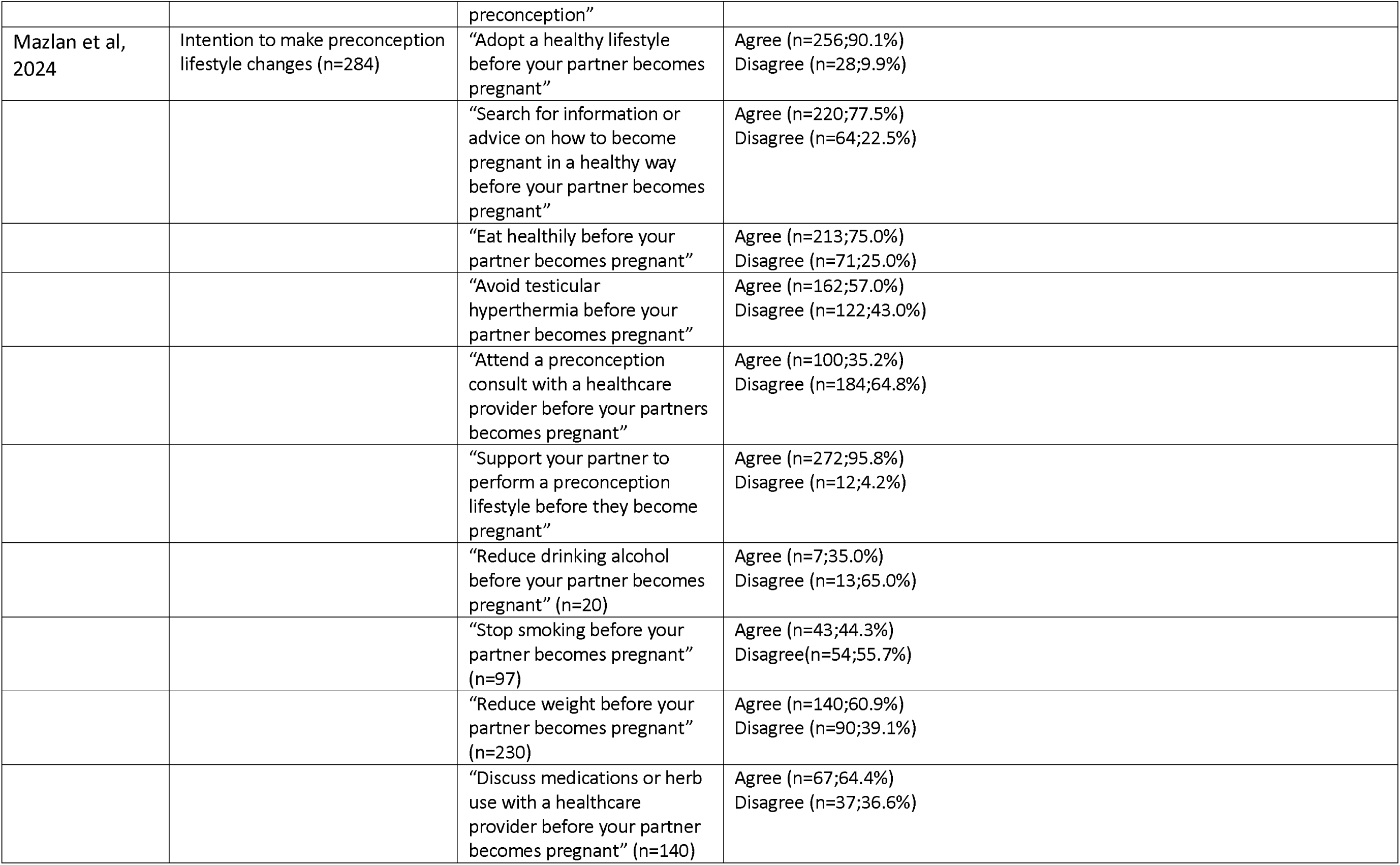
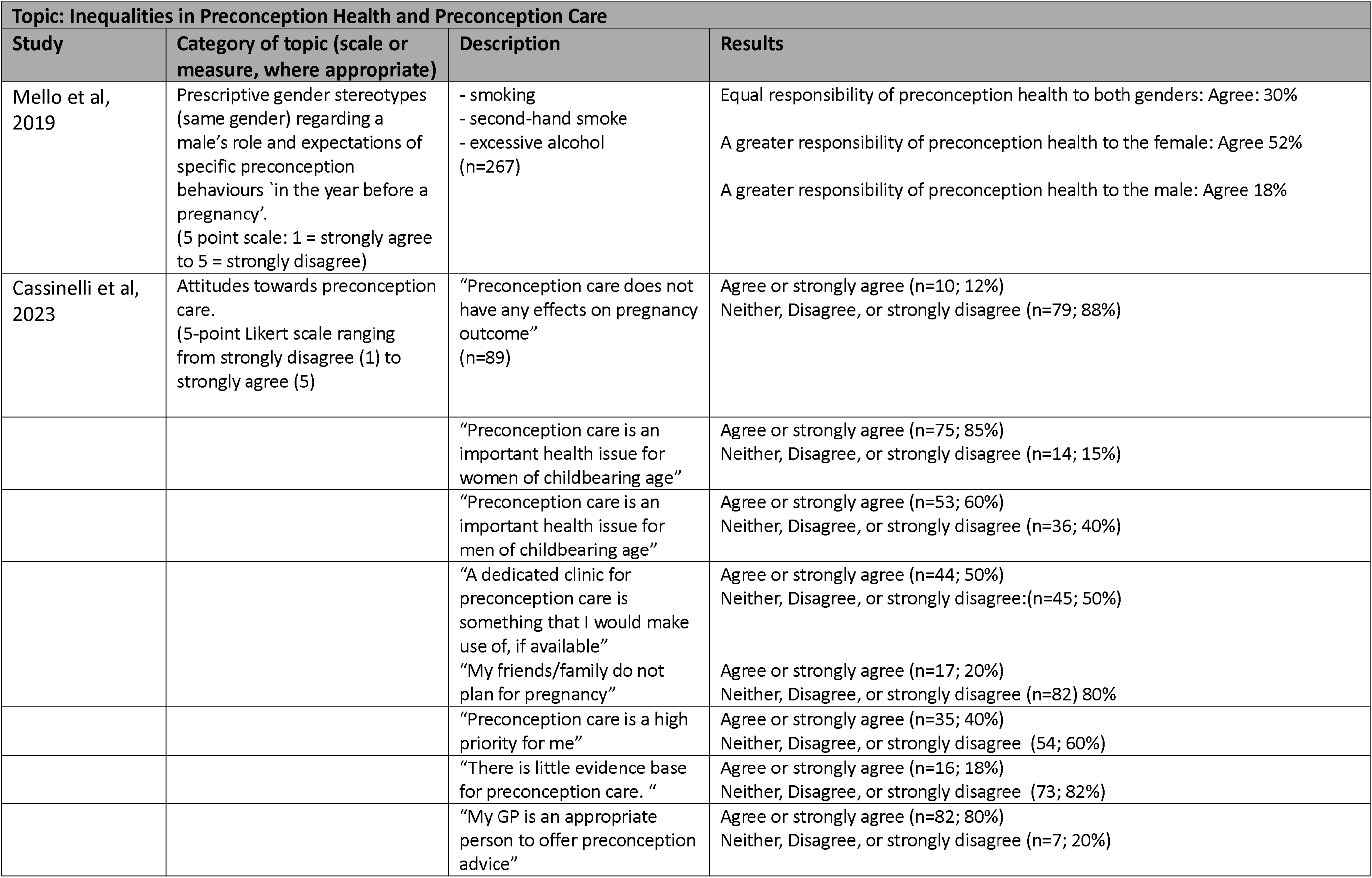
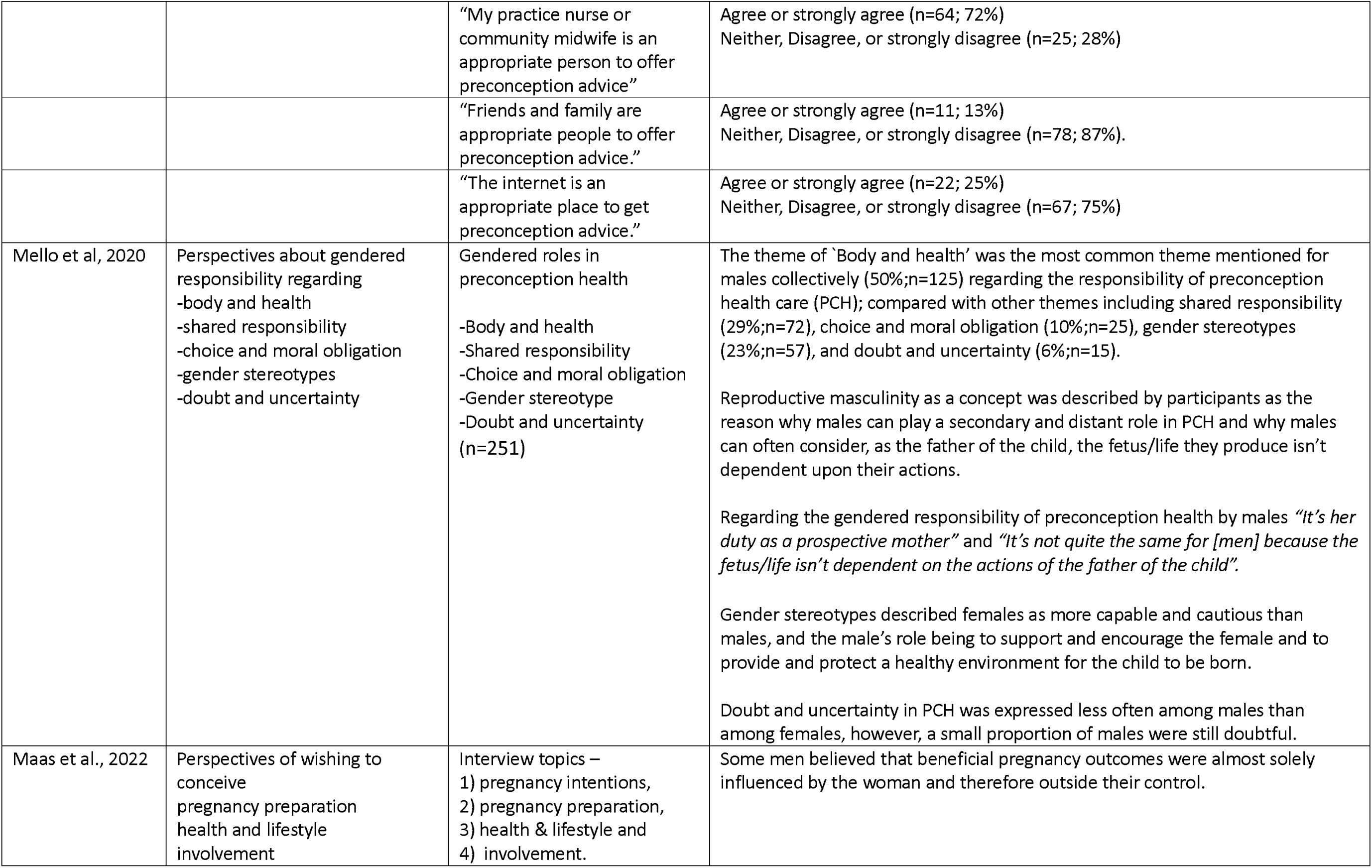
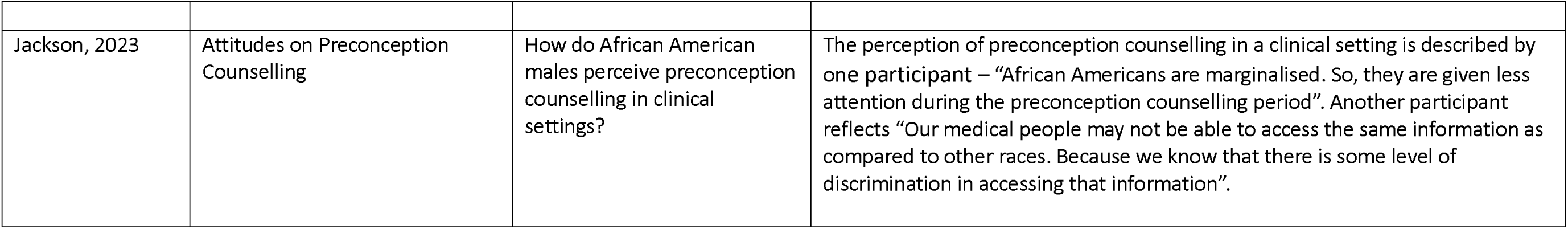
Summary table of findings from included studies, by theme.

### Importance of Preconception Health and Care

Four studies in this review reported the importance of preconception health and preconception care, as perceived by males aged 18 - 60 (See Table 2) (18, 19, 24, 25).

#### Preconception health

One cross-sectional study determined the existing awareness and beliefs of reproductive age adult males about reproductive life planning who were recruited from a student health centre of a large public university in the US (18). Results from this study report at least 50% of participants across different age groups believed it was important to understand the influence of family history when developing a reproductive life plan; <20 years (n=12;75%), 20-29 years (n=38;86%), for ≥ 30 years (n=4;57%) (p=0.03) (18). A high proportion of males from the aforementioned cross-sectional study (>85%), and another study which investigated prospective fathers perceptions regarding the requirement to engage in preconception care (25), agreed they made a contribution toward a healthy pregnancy and child. Although, for males considering a reproductive life plan, taking steps to optimise health before pregnancy was most often reported for males greater than 30 years old (85.7%) (18). The majority of prospective fathers (95%) agreed they can contribute to a healthy pregnancy, feel involved, and can influence the chances for the future of their child by living a healthy lifestyle (25).

#### Preconception care

The study conducted by Maas et al (2022) conducted in the Netherlands (25) revealed that from the prospective fathers who did not attend a preconception care consultation and were aware of the possibility for a preconception care consult (n=84), 65.5% did not opt for a preconception consultation with a health care provider as they believed it was not needed. A similar proportion of prospective fathers in the same circumstances did not attend a preconception consult but were aware; 63.1% did not opt for a preconception consultation as they agreed with the belief that they already knew enough about a healthy pregnancy. Further, based on qualitative findings from the same study, several men stated that available preconception care information was mainly communicated in a feminine tone of voice and narrative, suggesting a female cantered approach (25). From a study conducted in Ireland and the UK of adults of reproductive age and their beliefs and attitudes towards preconception care generally, one in two males (50%) agreed a dedicated clinic for preconception care is something they would use (19). The majority of participants in Ireland and the UK study agreed with the statement “My general practitioner [GP] is an appropriate person to offer preconception advice” (80%).

Qualitative evidence from a study conducted in the US (24) explored the attitudes of African American males toward preconception counselling. One participant from this study stated *“I just don’t feel comfortable, you know, talking about your feelings in and opening up to someone else”* while another participant revealed *“….Finding a professional that knows and is interested in helping and is also a good fit” “….. I need to know and trust people before I can ask for help and information”.* Another participant states “*No, I haven’t thought of going to a counsellor”….. but “It [preconception counselling] can help individuals to make an informed decision about their health, and the health of their future children which can lead to healthy pregnancy and outcomes and reduce risk of complications”.* These attitudes suggest a disconnect that males can experience when seeking support for matters related to preconception.

### Paternal Preconception Behaviours

Another key theme identified from the data was preconception health behaviours with relevant findings reported in six studies (See Table 2) (17, 19–22, 25).

A Belgian study (20) reported intention (self-efficacy) of males to make preconception lifestyle changes was associated with higher odds of adopting a healthy lifestyle, eat healthy, reduce alcohol consumption, or lose weight, however, confidence intervals were too wide to draw meaningful conclusions. Mixed-methods research from the Netherlands (25) surveyed prospective fathers exploring their preconception beliefs regarding preconception consultations. Contrary to other results in this review, which suggest that one in two males would use a dedicated preconception care clinic if made available, the mixed methods research from the Netherlands identified that many prospective fathers expressed they did not need a conversation with a health care provider as they already knew enough about a healthy pregnancy. One participant stated *“It [a GP proving PCC-information] is meant well, but I would think - I believe the doctor has better things to do than spending time on me to give me more information”.* Another participant declared *“Not drinking alcohol is part of the deal, I also don’t smoke so I didn’t have to quit, and I neither do drugs. I didn’t have to change much in that regard. I’m also not overweight, as far as I know. So I don’t have to adjust a lot, I thought it all will be fine.”*

A second study examined the preconception attitudes of Canadian couples toward the intention to eat healthy (21). This study reported longitudinal data for both affective attitudes (emotional based judgements about behaviour) and instrumental attitudes (perceived benefits and costs of the behaviour) to predict change regarding fruit and vegetable consumption and fat consumption from baseline (recruitment), 6 months from baseline, and 12 months from baseline. This study found affective attitudes were significant predictors for healthy eating among males (β= 0.55; p<0.01). The affective attitudes (β= −0.21; p=0.05) and instrumental attitudes (β= 0.25; p<0.05) of males were significant predictors of their fruit and vegetable consumption from baseline to twelve months only. Intention was a predictor of fruit and vegetable consumption 12 months from baseline (β = 0.09; p=0.05).

Research conducted in the US provided descriptive statistics to report the behavioural intentions of males before conception to avoid smoking cigarettes and avoid consuming excessive quantities of alcohol (17). Items were scored on a 5 point scale (1= strongly disagree to 5= strongly agree) and revealed that males place an emphasis on smoking avoidance (Mean 4.54:SD0.93) and to a lesser extent on too much alcohol (Mean 4.13:SD1.07) during preconception (17). This research also revealed current ‘every day’ male smokers were least likely to intend to engage in preconception health behaviours (β = −0.331; p<0.001;95% CI −0.551, – 0.111) followed by current ‘some days’ smokers (β = 0.165 p>0.05;95% CI −0.407, – 0.128) and former smokers (β = 0.049 p>0.05;95% CI −0.233, – 0.163), when compared with non-smokers (17).

A survey conducted in the Netherlands (25) among prospective fathers exploring their preconception beliefs regarding the cumulative effect of smoking and alcohol consumption on sperm quality found most study participants (88%) agreed these behaviours can affect the quality of their sperm. Quantitative research conducted in the UK found that more than half of males in the study (65%) agreed “It is important for men to consume a healthy preconception diet”. Approximately half (55%) of the males in the study agreed “It is important to for men to achieve a healthy weight during preconception” and “It is important for men to be physically active during preconception” (19).

A survey of married men conducted at a public health clinic in Malaysia reported their intention to make preconception lifestyle changes before their partner becomes pregnant (22). The majority of survey respondents (90%) intended to “adopt a healthy lifestyle before their partner becomes pregnant” and 75% of participants intended to “eat healthily before their partner becomes pregnant”. More than half of survey respondents (64%) intended to “discuss medications or herb use with a health care provider” and 61% intended to “reduce bodyweight before their partner becomes pregnant”. Only 35% of married men in this study intended to “attend a preconception consult with a healthcare provider before their partner becomes pregnant” or intended to “reduce drinking alcohol before their partner becomes pregnant”.

### Inequalities in Preconception Health and Preconception Care

Preconception health and care gender differences and ethnic disparities was the third key theme identified. Five studies in this review reported gender differences or ethnic disparities in preconception health and care (See Table 2) (17, 19, 23–25).

A study conducted in Ireland reported only 40% of males agreed that preconception care is a high priority for themselves, yet 85% of participants agreed with the statement that “preconception care is an important health issue for women of childbearing age” (19). An US study investigated same gender stereotypes regarding a male’s roles and expectations of preconception health behaviours by asking participants “….to think about whether [men/women] are responsible for engaging in certain activities in the year before pregnancy” (17). Approximately 30% of male survey panel members from this study agreed with an equal responsibility of preconception health to both genders (17).

Beliefs about gendered responsibility were also reported in a qualitative study conducted in the US (23). This study identified main themes based upon the examined personal and collective gender group responsibilities for preconception health of female and male respondents. The theme of ‘Body and health’ was the most common theme mentioned for males regarding the responsibility for optimal preconception health (50%); compared with other themes which included shared responsibility (29%), choice and moral obligation (10%), gender stereotypes (23%), and doubt and uncertainty (6%). The notion of reproductive masculinity was referred to in this study by male and female participants as a reason why males can play a secondary and distant role in preconception health and care and why they can often consider, as the prospective father of the child, *“It’s her duty as a prospective mother”* and *“It’s not quite the same for [men] because the fetus/life isn’t dependent on the actions of the father of the child”*.

At least one prospective father in the study conducted by Maas et al, 2002 (25) in the Netherlands believed beneficial pregnancy outcomes were almost solely influenced by the female and therefore mostly outside their control*. “I think it’s [contribution of one’s heath to that of the future child] for 80%-90% influenced by the women. So in the period that you try to conceive, you also try to live a little healthier. But after that, I think it mostly depends on the women.”*

The attitudes of African American males regarding preconception counselling (24) reveal *“African Americans are marginalised. So, they are given less attention during the preconception counselling period*”. Another participant reflects *“Our medical people may not be able to access the same information as compared to other races. Because we know that there is some level of discrimination in accessing that information”*.

## Discussion

This review describes the beliefs, attitudes and intentions of males regarding preconception health and care and pregnancy planning a reported in peer-reviewed literature. Nine relevant studies were identified, with only two deemed as high quality demonstrating the currently limited evidence base on this topic. The findings of this review can be summarised with reference to three overarching themes: 1) the importance of preconception health and preconception care; 2) paternal preconception behaviours; and 3) Inequalities in Preconception Health and Preconception Care.

A key finding of this review confirms males may not always consider their responsibilities during preconception or the ramifications of their preconception health (26) or feel comfortable consulting with a health professional about preconception. For males from certain ethnic groups, such as African Americans, they may even feel marginalised in contributing to pregnancy planning and preparation. As identified in some of the studies of this review, there appears to be a tendency for males to direct a greater level of responsibility to the female than to themselves regarding preconception health (for example – a male may place greater emphasis on their female partner avoiding smoking or eating healthy during preconception than for themselves). A tendency for males not to share the preconception responsibility with the female and in some cases feel marginalised during preconception care due to their race highlights preconception challenges that currently exist for paternal preconception care. Evidence supporting male preconception beliefs, attitudes, and intentions remains emergent yet is foundational to understanding male preconception behaviours and so is urgently required to understand males’ attitudes to adopt behaviour change.

Further findings highlights that males do not always opt for a preconception consultation and some hold the belief that they already know enough about a healthy pregnancy and therefore do not need a conversation with a health care provider. Preconception research outside this review which addresses paternal preconception knowledge reports similar findings; even when males believe preconception care is important, they are not always aware of their roles and responsibilities before pregnancy and therefore do not intend to consult with healthcare practitioners to optimise their health in preparation for pregnancy (26).

A male’s attitudes and intentions toward preconception consultations can be influenced by masculine stereotypes (23) which may consign undue strain and further challenge a male’s engagement with preconception health and care leading to disinterest or detachment when planning a pregnancy (23). In addition, practitioners often lack confidence to discuss preconception issues with their male patients (27). Many males may feel distant and secondary to the pregnancy planning process (28). This this appears to align with the fact they are often not included in preconception and pregnancy planning conversations in clinics (29, 30) nor often the focus of much preconception research (4).

Males’ belief that they already know enough about a healthy pregnancy and therefore do not need a conversation with a health care provider highlights the urgency of developing preconception health promotion specifically to males (31). Preconception policy and guidelines that directly help increase awareness among men are also required to address knowledge gaps and identify important areas of health behaviour change to support males in effectively contributing to reproductive health beyond fertility (32).

Another notable finding of this review is the understanding amongst males that smoking or drinking excessive amounts of alcohol in the period before pregnancy can adversely affect their sperm quality. This finding aligns with relevant research; alcohol abuse (33) and smoking (34) are associated with poor semen quality, and can influence adverse pregnancy and offspring outcomes (6). The negative health effects of smoking and excessive alcohol are serious public health issues (35, 36) which threaten reproductive health and yet may be effectively addressed by practitioners during preconception consultations. Interestingly, despite the identified understanding of males regarding smoking and alcohol consumption in this review, male current everyday smokers were least likely to intend to engage in preconception health behaviours (17). Smoking and excessive alcohol consumption which can be harmful to reproductive health are still identified behaviours undertaken during preconception by a substantial number of males preparing for fatherhood (37), this may be, in part at least, due to gaps in knowledge regarding preconception health amongst this subgroup of males (26). Our ability to target these males who continue smoking and/or excessive alcohol consumption during preconception requires further research to explore the experience and perspectives of this subgroup in more depth.

The influence of smoking and excessive alcohol consumption on male health during preconception is comprehensive and can have broader implications which extend beyond sperm quality and alter various aspects of preconception health, such as the consumption of a healthy diet or the maintenance of a healthy bodyweight, which in turn influence mental health (38, 39), and more broadly social and behavioural determinants of health such as social inclusion, employment, housing, sexual practices, or physical activity (40). This review identifies males’ attitudes regarding the adverse health effects of behaviours such as smoking and excessive alcohol during preconception, which is the first step to investigating the complexities of preconception behavioural intentions among males. Public health research initiatives aimed at reproductive health must consider further exploration and evaluation of paternal preconception beliefs and attitudes to help strengthen the current evidence base and effectively support the preconception behavioural intentions of males. Such research initiatives will help all providers better understand and support males’ participation in health behaviours before conception.

### Strengths and Limitations

This review is the first to summarise the current literature pertaining to the preconception behavioural beliefs, attitudes, and intentions specifically for males, it adheres to the PRISMA reporting guidelines and the review registration process, and is reinforced by a robust search strategy. However, such strengths must also be considered alongside limitations. Firstly, the review only included studies among generally healthy adult males and did not include males with a diagnosed illness, or disability, or males as health professionals or experts. Further, the studies included in the review were not often deemed good or high quality and some results identified in this review could not be utilised to draw any meaningful conclusions, despite being statistically significant (α>0.05), due to extremely wide confidence intervals. Only nine studies were included in this review so consequently the results reported are somewhat unvaried being narrowly focused, mostly sourced within cross-sectional data utilising small samples that report descriptive statistics from North America and Europe.

## Conclusion

This systematic review suggests the engagement and approach of many ales with regards to their important role in preconception health is far from optimal. Public health promotion, education, policy, guidelines and health services relating to preconception health that explicitly and specifically target males could improve understanding and awareness across all relevant providers and others in positions to better support males to engage in pregnancy planning and preconception health. Further research on the motivations behind paternal health beliefs and behaviours is needed to inform effective preconception health policy and interventions.

## Data Availability

All data extracted for this systematic review are presented as part of the manuscript.

## Declarations

## List of abbreviations

PROSPERO: Prospective Register of Systematic Reviews
PRISMA: Preferred Reporting Items for Systematic Reviews and Meta-Analyses
AMSTAR: Assessing the Methodological Quality of Systematic Reviews.
UTS: University of Technology Sydney
NOS: Newcastle Ottawa Scale
CASP: Critical Appraisal Skills Programme
GP: General Practitioner

## Ethics approval and consent to participate

Not applicable

## Consent for publication

Not applicable

## Availability of data and materials

All data extracted for this systematic review are presented as part of the manuscript.

## Author’s contributions

TC commenced the initial search strategy for the review, managed the study screening and data extraction processes, and was a major contributor to writing of the manuscript. This review was overseen by faculty supervisors AS, JA, & DS. All Authors read and approved the final manuscript.

## Acknowledgments

Not applicable.

## References

1. World Health Organization [WHO]. Preconception care: maximising the gains for maternal and child health - Policy brief World Health Organization - All rights reserved; 2013 15 February 2013. Contract No.: WHO REFERENCE NUMBER: WHO-FWC-MCA-13.02.

2. Hill B, Hall J, Currie S. Defining preconception: exploring the concept of a preconception population. BMC Pregnancy and Childbirth. 2020.

3. Harper T, Kuohung W, Sayres L, Willis MD, Wise LA. Optimizing preconception care and interventions for improved population health. Fertility and Sterility. 2023;120(3, Part 1):438–48.

4. Toivonen KI, Oinonen KA, Duchene KM. Preconception health behaviours: A scoping review. Preventive medicine. 2017;96:1–15.

5. Chivers BR, Boyle JA, Lang AY, Teede HJ, Moran LJ, Harrison CL. Preconception health and lifestyle behaviours of women planning a pregnancy: A cross-sectional study. Journal of Clinical Medicine. 2020;9(6).

6. Carter T, Schoenaker D, Adams J, Steel A. Paternal preconception modifiable risk factors for adverse pregnancy and offspring outcomes: a review of contemporary evidence from observational studies. BMC Public Health. 2023;23(1):509.

7. Manstead ASR, Parker D. Evaluating and extending the theory of planned behaviour. European review of social psychology. 1995;6(1):69–95.

8. Sur MH, Jung J, Shapiro DR. Theory of planned behavior to promote physical activity of adults with physical disabilities: Meta-analytic structural equation modeling. Disabil Health J. 2022;15(1):101199.

9. McDermott MS, Oliver M, Simnadis T, Beck EJ, Coltman T, Iverson D, et al. The Theory of Planned Behaviour and dietary patterns: A systematic review and meta-analysis. Preventive Medicine. 2015;81:150–6.

10. Robbins CL, Gavin L, Carter MW, Moskosky SB. The link between reproductive life plan assessment and provision of preconception care at publicly funded health centers. Perspectives on sexual and reproductive health. 2017;49(3):167–72.

11. Page MJ, McKenzie JE, Bossuyt PM, Boutron I, Hoffmann TC, Mulrow CD, et al. The PRISMA 2020 statement: An updated guideline for reporting systematic reviews. PLOS Medicine. 2021;18(3):e1003583.

12. Shea BJ, Reeves BC, Wells G, Thuku M, Hamel C, Moran J, et al. AMSTAR 2: a critical appraisal tool for systematic reviews that include randomised or non-randomised studies of healthcare interventions, or both. Bmj. 2017;358:j4008.

13. Covidence. Systematic review software Melbourne, Australia Veritas Health Innovation; [Available from: www.covidence.org.

14. Stang A. Critical evaluation of the Newcastle-Ottawa scale for the assessment of the quality of nonrandomized studies in meta-analyses. European journal of epidemiology. 2010;25:603–5.

15. Sharmin S, Kypri K, Khanam M, Wadolowski M, Bruno R, Mattick RP. Parental Supply of Alcohol in Childhood and Risky Drinking in Adolescence: Systematic Review and Meta-Analysis. International Journal of Environmental Research and Public Health. 2017;14(3).

16. Lockwood C, Munn Z, Porritt K. Qualitative research synthesis: methodological guidance for systematic reviewers utilizing meta-aggregation. Int J Evid Based Healthc. 2015;13(3):179–87.

17. Mello S, Tan ASL, Sanders-Jackson A, Bigman CA. Gender Stereotypes and Preconception Health: Men’s and Women’s Expectations of Responsibility and Intentions to Engage in Preventive Behaviors. Maternal & Child Health Journal. 2019;23(4):459–69.

18. Kransdorf LN, Raghu TS, Kling JM, David PS, Vegunta S, Knatz J, et al. Reproductive life planning: a cross-sectional study of what college students know and believe. Maternal and child health journal. 2016;20:1161–9.

19. Cassinelli EH, McClure A, Cairns B, Griffin S, Walton J, McKinley MC, et al. Exploring Health Behaviours, Attitudes and Beliefs of Women and Men during the Preconception and Interconception Periods: A Cross-Sectional Study of Adults on the Island of Ireland. Nutrients. 2023;15(17).

20. Goossens J, Van Hecke A, Beeckman D, Verhaeghe S. The intention to make preconception lifestyle changes in men: Associated socio-demographic and psychosocial factors. Midwifery. 2019;73:8–16.

21. Bassett-Gunter RL, Levy-Milne R, Naylor PJ, Symons Downs D, Benoit C, Warburton DER, et al. Oh baby! Motivation for healthy eating during parenthood transitions: a longitudinal examination with a theory of planned behavior perspective. International Journal of Behavioral Nutrition and Physical Activity. 2013;10:1–11.

22. Mazlan SSA, Ismail IZ, Devaraj NK. Intention to make preconception lifestyle changes among married men in an urban primary care clinic and its association with self-efficacy. Journal of Men’s Health. 2024;20(11):112–9.

23. Mello S, Stifano S, Tan AS, Sanders-Jackson A, Bigman CA. Gendered conceptions of preconception health: a thematic analysis of men’s and women’s beliefs about responsibility for preconception health behavior. Journal of Health Communication. 2020;25(5):374–84.

24. Jackson ND. Exploring African American Male Attitudes on Preconception Counseling. 2023.

25. Maas VYF, Poels M, Stam AL, Lieftink N, Franx A, Koster MPH. Exploring male perceptions regarding the need to engage in preconception care–a mixed-method study amongst Dutch (prospective) fathers. The European Journal of Contraception & Reproductive Health Care. 2022;27(4):322–9.

26. Rabiei Z, Shariati M, Mogharabian N, Tahmasebi R, Ghiasi A, Motaghi Z. Men’s knowledge of preconception health: A systematic review. J Family Med Prim Care. 2023;12(2):201–7.

27. Hogg K, Rizio T, Manocha R, McLachlan RI, Hammarberg K. Men’s preconception health care in Australian general practice: GPs’ knowledge, attitudes and behaviours. Aust J Prim Health. 2019;25(4):353–8.

28. Kotelchuck M. The impact of father’s health on reproductive and infant health and development. Engaged Fatherhood for Men, Families and Gender Equality. 2022:31.

29. Withanage NN, Botfield JR, Srinivasan S, Black KI, Mazza D. Effectiveness of preconception interventions in primary care: a systematic review. British Journal of General Practice. 2022;72(725):E865–E72.

30. Dorney E, Boyle JA, Walker R, Hammarberg K, Musgrave L, Schoenaker D, et al. A Systematic Review of Clinical Guidelines for Preconception Care. Semin Reprod Med. 2022.

31. Shawe J, Patel D, Joy M, Howden B, Barrett G, Stephenson J. Preparation for fatherhood: a survey of men’s preconception health knowledge and behaviour in England. PLoS One. 2019;14(3):e0213897.

32. Kotelchuck M, Lu M. Father’s role in preconception health. Maternal and child health journal. 2017;21(11):2025–39.

33. Finelli R, Mottola F, Agarwal A. Impact of Alcohol Consumption on Male Fertility Potential: A Narrative Review. Int J Environ Res Public Health. 2021;19(1).

34. Bundhun PK, Janoo G, Bhurtu A, Teeluck AR, Soogund MZS, Pursun M, et al. Tobacco smoking and semen quality in infertile males: a systematic review and meta-analysis. BMC Public Health. 2019;19(1):36.

35. Holland L, Reid N, Hewlett N, Toombs M, Elisara T, Thomson A, et al. Alcohol use in Australia: countering harm with healing. The Lancet Regional Health – Western Pacific. 2023;37.

36. Dai X, Gil GF, Reitsma MB, Ahmad NS, Anderson JA, Bisignano C, et al. Health effects associated with smoking: a Burden of Proof study. Nature Medicine. 2022;28(10):2045–55.

37. Shawe J, Patel D, Joy M, Howden B, Barrett G, Stephenson J. Preparation for fatherhood: A survey of men’s preconception health knowledge and behaviour in England. PLoS ONE. 2019;14(3).

38. Ning K, Gondek D, Patalay P, Ploubidis GB. The association between early life mental health and alcohol use behaviours in adulthood: A systematic review. PLOS ONE. 2020;15(2):e0228667.

39. Plurphanswat N, Kaestner R, Rodu B. The effect of smoking on mental health. American journal of health behavior. 2017;41(4):471–83.

40. Australian Institute of Health & Welfare A. Social determinants of health 2022 [Available from: https://www.aihw.gov.au/reports/australias-health/social-determinants-of-health#Social%20inclusion.

