## Supplementary Table 2 for "The health beliefs, attitudes, and intentions of males toward pregnancy planning and preconception health and care: a systematic review"

Supplementary Table 2 – Quality Assessment: The Critical Appraisal Skills Programme [CASP]

| **The Critical Appraisal Skills Programme [CASP] (Qualitative) - Criteria** | | | | | | | | | | | | | | | | | | | | | | | | | | | |  |
| --- | --- | --- | --- | --- | --- | --- | --- | --- | --- | --- | --- | --- | --- | --- | --- | --- | --- | --- | --- | --- | --- | --- | --- | --- | --- | --- | --- | --- |
| **First author & Year** | **Section A** | | | | | | | | | | | | | | | | | | **Section B** | | | | | | | | | **Section C** |
|  | **Was there a clear statement of research aims?** | | | **Is the method appropriate?** | | | **Was the research design appropriate?** | | | **Was the recruitment strategy appropriate?** | | | **Was data collected in a way that addressed the research issue?** | | | **Has the relationship between researcher and participants been adequately considered?** | | | **Have ethical issues been considered?** | | | **Was the data analysis rigorous?** | | | **Is there a clear statement of findings?** | | | **How valuable is the research?** |
|  | Yes | ? | No | Yes | ? | No | Yes | ? | No | Yes | ? | No | Yes | ? | No | Yes | ? | No | Yes | ? | No | Yes | ? | No | Yes | ? | No |  |
| Maas et al., 2022 | ☒ |  |  | ☒ |  |  | ☒ |  |  | ☒ |  |  | ☒ |  |  |  | ☒ |  | ☒ |  |  | ☒ |  |  | ☒ |  |  |  |
| *Notes* | A clear aim is presented | | | Exploring male perceptions | | | The research design was justified | | | Participants recruited following survey completion | | | Data collection methods were adequate | | | Participant consent was checked but researchers did not consider their influence in formation of the research question. | | | A `Medical Ethical Review Board’ has approved the study | | | Thematic analysis conducted and clearly explained. | | |  | | | **High value** Researchers identify the contribution this research makes to the field and the opportunities for future research arising from the study findings. |
|  | Yes | ? | No | Yes | ? | No | Yes | ? | No | Yes | ? | No | Yes | ? | No | Yes | ? | No | Yes | ? | No | Yes | ? | No | Yes | ? | No |  |
| Mello et al., 2020 | ☒ |  |  | ☒ |  |  |  | ☒ |  | ☒ |  |  |  | ☒ |  | ☒ |  |  | ☒ |  |  | ☒ |  |  |  | ☒ |  |  |
| *Notes* | Research question/s clearly described. | | | Assessing the beliefs of participants | | | The research design justification was not clear | | | Recruitment clearly explained | | | The qualitative methodology was not justified nor clearly explained. Also, open text survey responses do not provide rich and meaningful qualitative data. | | | The relationship between researchers and participants are covered. | | | The Northeastern University's institutional review board approved the study | | | Thematic analysis conducted and explained | | | Results in this study are aggregated which is difficult to report for males only. | | | **Moderate value** Researchers do not identify the contrition this research makes to the field and opportunities for future research. Also, the discussion focused mainly on women not men. |
|  | Yes | ? | No | Yes | ? | No | Yes | ? | No | Yes | ? | No | Yes | ? | No | Yes | ? | No | Yes | ? | No | Yes | ? | No | Yes | ? | No |  |
| Jackson, 2023 | ☒ |  |  | ☒ |  |  | ☒ |  |  | ☒ |  |  | ☒ |  |  | ☒ |  |  | ☒ |  |  | ☒ |  |  | ☒ |  |  |  |
| *Notes* | Research question/s clearly described. | | | Assessing the attitudes of participants | | | The research design justification was clear | | | Recruitment clearly explained | | | Data collection methods were adequate | | | The relationship between researchers and participants are covered. | | | The Northcentral University's institutional review board approved the study | | | Thematic analysis conducted and explained | | |  | | | **High value** Dissertation highlights the preconception opportunities of an ethnic minority group and makes recommendations for future research. |
