## Supplementary File 1 for "The health beliefs, attitudes, and intentions of males toward pregnancy planning and preconception health and care: a systematic review"

Male preconception beliefs, attitudes, and intentions – Search Strategy

1) **Medline (OVID)**

**Controlled vocabulary – Medical subject headings [MESH]**

^mp – multi-purpose

^sh – MesH Subject Heading

| # | Query from 3^rd^ January, 2024 |
| --- | --- |
| S1 | (preconception or pre-conception or periconception or peri-conception or prepregnancy or pre-pregnancy or interconception or pregnan* plan* or inten* pregnan*).mp. |
| S2 | Preconception care .sh. |
| S3 | S1 OR S2 |
| S4 | (Male* or father* or husband* or paternal or men’s health).mp. |
| S5 | (Male or Fathers or Men’s health) .sh. |
| S6 | S4 OR S5 |
| S7 | (health adj1 belief*) or (health adj1 attitude*) or (health adj1 intention*) or (health adj1 acceptance*) or (health adj1 thought*) or (health adj1 viewpoint*) or (health adj1 perspective*) or (health adj1 perception*) or (health adj1 outlook*) or (health adj1 approach*) or (health adj1 opinion*) .mp. |
| S8 | (Attitude to health or Intention or Health, Knowledge, Attitudes, Practice).sh. |
| S9 | S7 OR S8 |
| S10 | S3 AND S6 AND S9 |

*Truncation

2) **Embase (OVID)**

**Controlled vocabulary – Science Thesaurus [Emtree]**

^mp – multi-purpose

^sh – Subject Headings

| # | Query from 3^rd^ January, 2024 |
| --- | --- |
| S1 | (Preconception or pre-conception or periconception or peri-conception or prepregnancy or pre-pregnancy or interconception or pregnan* plan* or inten* pregnan*) .mp. |
| S2 | Prepregnancy care .sh. |
| S3 | S1 OR S2 |
| S4 | (Male* or father* or husband* or paternal or men’s health) [^mp] |
| S5 | (Male or Father or Husband or Men’s health) .sh. |
| S6 | S4 OR S5 |
| S7 | (health adj1 belief*) or (health adj1 attitude*) or (health adj1 intention*) or (health adj1 acceptance*) or (health adj1 thought*) or (health adj1 viewpoint*) or (health adj1 perspective*) or (health adj1 perception*) or (health adj1 outlook*) or (health adj1 approach*) or (health adj1 opinion*) [^mp] |
| S8 | (Health belief or Attitude to health or Attitude to pregnancy or Paternal attitude).sh. |
| S9 | S7 OR S8 |
| S10 | S3 AND S6 AND S9 |

*Truncation

3) **PubMed**

**Controlled vocabulary – Medical subject headings [MESH].**

^af – All fields

^sh – MeSH Terms

| # | Query from 3^rd^ January, 2024 |
| --- | --- |
| S1 | (Preconception or pre-conception or periconception or peri-conception or prepregnancy or pre-pregnancy or interconception or pregnan plan* or inten* pregnan*).af. |
| S2 | Preconception care .sh. |
| S3 | S1 OR S2 |
| S4 | (Male* or father* or husband* or paternal or men’s health) .af. |
| S5 | (Male or men or fathers or paternal behavior) .sh. |
| S6 | S4 OR S5 |
| S7 | “health belief” [Title/Abstract: ~1] or “health attitude” [Title/Abstract: ~1] or “health intention” [Title/Abstract: ~1] or “health acceptance” [Title/Abstract: ~1] or “health thought” [Title/Abstract: ~1] or “health viewpoint” [Title/Abstract: ~1] or “health perspective” [Title/Abstract: ~1] or “health perception” [Title/Abstract: ~1] or “health outlook” [Title/Abstract: ~1] or “health approach” [Title/Abstract: ~1] or “health opinion” [Title/Abstract: ~1] |
| S8 | (Attitude to health or health belief model or intention) .sh. |
| S9 | S7 OR S8 |
| S10 | S3 AND S6 AND S9 |

*Truncation

4) **CINAHL (EBSCO)**

**Controlled vocabulary – Subject headings [subject headings authority profile]**

^TX – all text

^MM – Major subject heading

| # | Query from 3^rd^ January, 2024 |
| --- | --- |
| S1 | (Preconception or pre-conception or periconception or peri-conception or prepregnancy or pre-pregnancy or interconception or pregnan* plan* or inten* pregnan*). TX. or (Prepregnancy care).MM. |
| S2 | Male* or father* or husband* or paternal or men’s health).TX. or (Male or men or fathers or men`s health or paternal role).MM. |
| S3 | (health W1 belief*) or (health W1 attitude*) or (health W1 intention*) or (health W1 acceptance*) or (health W1 thought*) or (health W1 viewpoint*) or (health W1 perspective*) or (health W1 perception*) or (health W1 outlook*) or (health W1 approach*) or (health W1 opinion*).TX. |
| S4 | (Health beliefs or intention or paternal attitudes or parental attitudes or attitude to pregnancy or social attitudes).MM. |
| S5 | S3 OR S4 |
| S6 | S1 AND S2 AND S5 |

*Truncation

5) **APA PsycINFO (EBSCO)**

**Controlled vocabulary – APA Thesaurus of Psychological Index Terms**

^TX – all text

^MA – MeSH subject heading

| # | Query from 3^rd^ January, 2024 |
| --- | --- |
| S1 | (Preconception or pre-conception or periconception or peri-conception or prepregnancy or pre-pregnancy or interconception or pregnan* plan* or inten* pregnan*).TX. |
| S2 | (Male* or father* or husband* or paternal or men’s health).TX. |
| S3 | (health W1 belief*) or (health W1 attitude*) or (health W1 intention*) or (health W1 acceptance*) or (health W1 thought*) or (health W1 viewpoint*) or (health W1 perspective*) or (health W1 perception*) or (health W1 outlook*) or (health W1 approach*) or (health W1 opinion*).TX. |
| S4 | (Male attitudes or attitudes or attitude formation or attitude measures or intention) .MA. |
| S5 | S3 OR S4 |
| S6 | S1 AND S2 AND S5 |

*Truncation

6) **Scopus**

**Controlled vocabulary – *Note Scopus does not use a controlled vocabulary/ subject heading.**

^– Search within Article title, Abstract, Keywords

| # | Query from 3^rd^ January, 2024 |
| --- | --- |
| S1 | (Preconception or pre-conception or periconception or peri-conception or prepregnancy or pre-pregnancy or interconception or pregnan* W/1 plan* or inten* W/1 pregnan*) ^ |
| S2 | (Male* or father* or husband* or paternal or men’s health) ^ |
| S3 | (health W/1 belief*) or (health W/1 attitude*) or (health W/1 intention*) or (health W/1 acceptance*) or (health W/1 thought*) or (health W/1 viewpoint*) or (health W/1 perspective*) or (health W/1 perception*) or (health W/1 outlook*) or (health W/1 approach*) or (health W/1 opinion*) ^ |
| S4 | S1 AND 2 AND 3 |

*Truncation

7) **ISI Proceedings/Web of Science**

**Controlled vocabulary – *Note ISI Proceedings does not use a controlled vocabulary/ subject heading.**

^af – All fields

| # | Query from 3^rd^ January, 2024 |
| --- | --- |
| S1 | (Preconception or pre-conception or periconception or peri-conception or prepregnancy or pre-pregnancy or interconception or pregnan* plan* or inten* pregnan*) .af. |
| S2 | (Male* or father* or husband* or paternal or men’s health) .af. |
| S3 | (health belief*) or (health attitude*) or (health intention*) or (health acceptance*) or (health thought*) or (health viewpoint*) or (health perspective*) or (health perception*) or (health outlook*) or (health approach*) or (health opinion*) .af. |
| S4 | S1 AND S2 AND S3 |

*Truncation
