## Supplementary Table 1 for "The health beliefs, attitudes, and intentions of males toward pregnancy planning and preconception health and care: a systematic review"

Supplementary Table 1 – Quality Assessment: The Newcastle Ottawa Scale [NOS]

| **Newcastle-Ottawa Critical Analysis (Cross-sectional) - Criteria** | | | | | | | | | |
| --- | --- | --- | --- | --- | --- | --- | --- | --- | --- |
| **First author & Year** | **SELECTION*** | | | | **COMPARABILITY**** | **OUTCOME***** | | **TOTAL** | **Quality** |
|  | **Representativeness of the sample** | **Sample size** | **Non-Response rate** | **Ascertainment of the exposure^** | **Potential confounders were investigated** | **Assessment of the outcome ^^** | **Statistical test** |  |  |
| Mello et al, 2019 | ☒ |  |  | ☒ | ☒ | ☒ | ☒ | 5 | Fair |
| Kransdorf et al., 2016 |  |  |  | ☒ | ☒ | ☒ | ☒ | 4 | Fair |
| Maas, 2022 | ☒ |  |  | ☒ | ☒ | ☒ | ☒ | 5 | Fair |
| Cassinelli et al., 2023 | ☒ | ☒ |  | ☒ |  |  | ☒ | 4 | Fair |
| Goossens et al., 2019 |  |  |  |  | ☒ | ☒ | ☒ | 3 | Poor |
| Bassett-Gunter et al., 2013 | ☒ |  |  |  | ☒ | ☒ | ☒ | 4 | Fair |
| Mazlan et al., 2024 | ☒ |  |  | ☒☒ | ☒ | ☒ | ☒ | 6 | Fair |

*A maximum of five (5) stars can be awarded for the selection domain

^A maximum of two stars (2) can be awarded for the ascertainment of exposure

** A maximum of two (2) stars can be awarded for the comparability domain

*** A maximum of three (3) stars can be awarded for the outcome domain

^^ A maximum of two (2) stars can be awarded for the ascertainment of outcome
